## Supplementary 1: Systematic Narrative Review Methodology and Versions of Health System Research Frameworks (Version 0 to 3) for "The Dialogic Health Systems Research Framework (DHSRF): A tool for facilitating self-criticality, researcher interactions and knowledge management in Health Systems Research & Policy Studies"

**Supplementary 1: Systematic Narrative Review Methodology**
**and Versions of Health System Research Frameworks (Version 0 to 3)**

### Systematic Narrative Review Methodology

The first phase of this project involved conducting a systematic narrative review. The primary objective was to review the existing literature and map the various health systems frameworks widely used in health systems research and policy studies (HSR&PS) globally. The overall review objective was to make futuristic recommendations. Hence, the systematic narrative review was located within the discourse on frameworks used in health systems research in the last two decades, from 2000 to 2022.

The second objective was to develop a Health Systems *Research* Framework by examining the strengths and limitations of the reviewed health systems frameworks, guided by Alma Ata's (1978) PHC approach and the complex adaptive systems lens.

The following two objectives were framed for phase 1 -

Table 1.1: Research Objectives and Research Question for the Systematic Review

| | **Research Objective** | **Research Question** |
|---|---|---|
| **1** | A systematic narrative review of health systems frameworks from the year 2000 onwards | "What are the various frameworks used in the contemporary discourse of health systems research and policy studies from 2000 onwards?" |
| **2** | Developing a framework to fill the gaps evident through the review of health system frameworks with reference to the PHC approach of Alma Ata (1978) and the complex adaptive systems lens | a) What elements of the PHC approach of Alma Ata 1978 and the complex adaptive systems lens are not conceptualized or adopted in practice in available HSR&PS?<br><br>b) How can they be incorporated to develop a more holistic framework? |

#### Search Strategy and Inclusion Criteria

The first research question informed the systematic search for literature on the subject. We developed three inclusion criteria and four exclusion criteria to meet the above-mentioned research questions. We were only interested in studies on health systems frameworks. So irrespective of disciplines, we included any peer reviewed journal publications, reports, commentary, position papers that conceptualized, defined, reported and/or applied a health systems framework; that which assessed or evaluated whole health systems (and not just any components or aspects of health systems); that which explicitly stated their health systems framework. Table 1.2 shows the inclusion and exclusion criteria which were used to select papers.

Table 1.2: Inclusion & exclusion criteria used to select papers for the research questions

| **Research Question** | **Inclusion & Exclusion Criteria** |
|---|---|
| "What are the various frameworks used in the contemporary discourse of health systems research and policy studies from 2000 onwards?" | **Inclusion Criteria**<br><br>● Documents (journal articles, reports, commentary, position papers) that conceptualize, define, report and/or apply a health systems framework<br>● Documents (journal articles, reports, commentary, position papers) that assess or evaluate whole health systems and not just any components or aspects of health systems<br>● Documents (journal articles, reports, commentary, position papers) that explicitly state their health systems framework<br><br>**Exclusion Criteria**<br><br>● Documents (journal articles, reports, commentary, position papers) that do not conceptualize, define, report and/or apply a health systems framework<br>● Documents (journal articles, reports, commentary, position papers) that do not assess or evaluate whole health systems<br>● Documents (journal articles, reports, commentary, position papers) that do not explicitly state their health systems framework.<br>● Documents (journal articles, reports, commentary, position papers) that are reviews of health systems research frameworks |

The search of the papers was limited to full-text English language documents between 2000 and 2022 time period accessible through the databases subscribed by the Jawaharlal Nehru University (JNU) library.

We searched four electronic databases (ScienceDirect, Pubmed, Ebscohost, Proquest) using three keywords ("Health") ("Systems") ("Framework") combined with boolean operator

(AND). For example, the keyword *health* was combined with the word *systems* and with the word *framework*. Relating terms of each of the keywords were not used for searching the studies as we were only interested to review studies which include any health systems framework.

The keywords were first searched in titles of each of the databases using the boolean operator (AND) and later were screened by abstract. A similar search strategy was used for all three databases. Except for ScienceDirect database where the database does not give any provision to apply the boolean operator (AND). Thus, for this database all three keywords were combined. For example, all three keywords were combined "Health Systems Framework" and searched in the title of the database. Articles were screened by reviewing the abstract after screening the articles in the titles from each of the four databases. We searched all four databases simultaneously to reduce the biases in the selection of the articles. Search strategies used in each of the databases including the keywords and search sets are outlined in Table 1.3

Table 1.3: The search strategy across four databases

| **Search Strategy** | | **ScienceDirect Database** | **Pubmed Database** | **EBSCOhost Database** | **Proquest Database** |
|---|---|---|---|---|---|
| Keywords | 1. | Health | Health | Health | Health |
| | 2. | Systems | Systems | Systems | Systems |
| | 3. | Framework | Framework | Framework | Framework |
| Search Set | | "Health Systems Frameworks" | 1 AND 2 AND 3 | 1 AND 2 AND 3 | 1 AND 2 AND 3 |
| Total number of studies identified from each database | | 89 | 177 | 1351 | 168 |
| Total number of studies screened by title and abstract from each database | | 43 | 129 | 203 | 55 |

Two researchers independently screened the studies. Each of the two researchers was assigned two databases. To reduce biases in the screening of the studies all the databases were searched simultaneously by the two researchers. After screening the studies by abstract from each database, the final screening of the articles was done in consultation with a third reviewer to reduce the biases in the final selection of the studies.

We included peer reviewed journal publications— any type of report, commentary, position papers that conceptualized health systems framework or defined health systems framework; reported and/or applied a health systems framework; assessed or evaluated health systems frameworks in their studies. Articles were searched for all disciplines, including any type of studies. Thus, screening of the articles was done purposively based on the research questions.

The review identified from the four databases includes a search of a total of 1785 studies of which 430 studies were screened by title and abstract. After deleting 87 duplicate records across the four databases, 343 studies were sought for full access. On 256 studies, inclusion and exclusion criteria were applied and based on non-eligibility, 196 studies were excluded. Thus, a total of 60 studies were selected for analysis of health systems frameworks. The study selection procedure and its results are illustrated in Figure 1.

Figure 1: Flow Diagram of Study Selection Procedure and Results

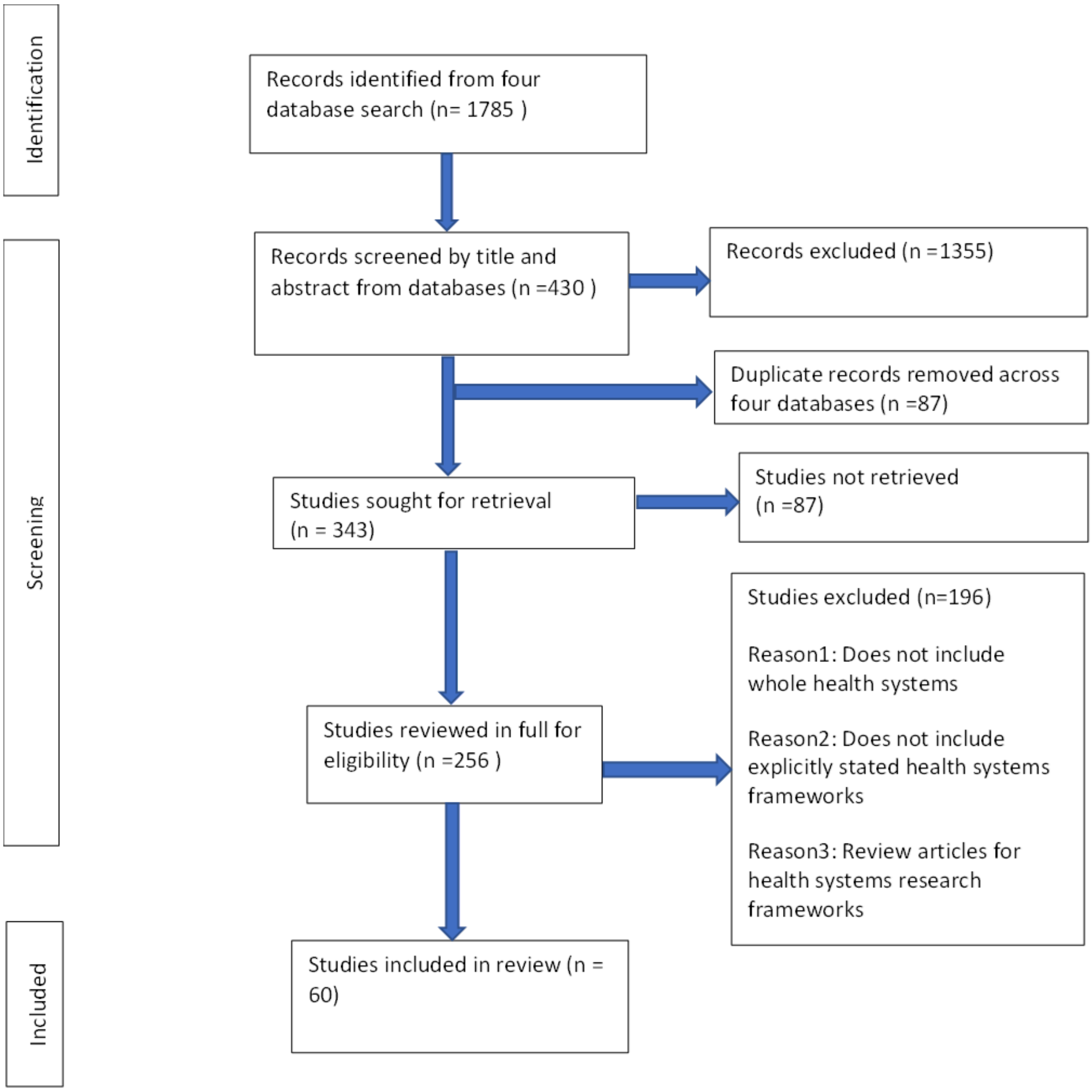

Flow diagram of study selection procedure and results (adapted from PRISMA 2020)

**Assessment of the quality of included studies**

We assessed the quality of 47 studies out of the selected set of 60 studies using a published mixed method tool (Hong et al. 2018). We could not appraise the quality of 13 studies which did not use any methods for describing and conceptualizing the frameworks. The mixed methods appraisal tool (MMAT) provided the criteria to appraise the quality of the diverse studies using quantitative and qualitative research designs describing the health systems frameworks. Forty-seven studies provided adequate descriptions for assessing the quality of studies such as research objectives, methods, study participants and research process. Each of the 47 studies was appraised using the user guide and rated as high, medium and low quality.

**Development of the analytical framework for analysis of the selected 60 studies:**

The Primary Health Care (PHC) approach enshrined in the Alma Ata Declaration of 1978, presents one of the most comprehensive conceptualisations of health systems so far. The PHC approach articulated in the declaration and scholarship that critically engaged and expanded the ideas encompassed in the spirit of the approach informed the development of the initial conceptual and methodological configuration of the health systems framework used in the present study (Labonte, Sanders, Packer & Schaay, 2017; Priya et al., 2019; Rakhal et al., 2019; Ghodajkar et al., 2019; Mathpati, Payyappallimana, Shankar & Porter, 2022). Informed by the health systems design of the PHC declaration and subsequent conceptual development in this area, the PHC approach was applied in the research project involving the whole health system, not to primary-level services and social determinants of health alone. It was further informed by the complexity theory which represents newer advances in HSR through its conceptualisation of typical attributes of complex systems. This was used to develop the initial analytical framework for the systematic narrative review. The initial framework is called Health Systems Research Framework Version 0 (HSRF-0).

Health Systems Research Framework Version 0 (HSRF-0)

**Analytical details of the selected studies**

| **Name of study** | | | **Observations** |
|---|---|---|---|
| **Conceptualisation of Health Systems** | **Boundary** | | |
| | **Elements of the Health System and Its Context** | Ecosystem | |
| | | Socio-political Contexts | |
| | | Meaning System | |
| | | Health Care System (Societal elements other than health health service system | |
| | **Relationship between elements** | | |
| **Methodological Approaches** | **Analytical Approaches** | Historical analysis | |
| | | Political Economy approach | |
| | | Analysis of Power and Hierarchy | |
| | | Epidemiological Orientation | |

| | | |
|---|---|---|
| | | Knowledge Pluralism |
| | | Health System Vantage Point |
| | **Operational Approaches** | |
| **Recommendations** | | |
| **Strengths** | | |
| **Limitations** | | |
| **Research Paradigm** | | |

In addition, the following details for each of the selected studies were also obtained for analysis.

**Descriptive details of the selected studies**

| |
|---|
| **Name of the study** |
| **Author/s** |
| **Year of Publication** |
| **Location (study site, if any)** |
| **Funding** |
| **Type of Research** |
| **Methodology of developing the framework** |
| **Structure of Disciplinary Interaction in the Study (Multi/Trans/Interdisciplinary)** |
| **Aim and Purpose of the study** |
| **Level of Analysis(Global/National/State/District)** |
| **Quality Of the Paper - Category (High/Medium/Low)** |

**Data extraction sheet**

Please click on the following link

**HSF Summary Table Final.xlsx**

- **Versions of Health System Research Frameworks (Version 0 to 3)**

This section presents the four different iterations of the Health System Research Frameworks as they evolved during the research process including the final one, the Dialogic Health Systems Research Framework.

**The Health Systems Research Framework Version - 0**

1. Analytical details of the selected studies

<table>
<tr><td>Name of study</td><td colspan="2"></td><td>Observations</td></tr>
<tr><td rowspan="7">Conceptualisation of Health Systems</td><td colspan="2">Boundary</td><td></td></tr>
<tr><td rowspan="4">Elements of the Health System and Its Context</td><td>Ecosystem</td><td></td></tr>
<tr><td>Socio-political Contexts</td><td></td></tr>
<tr><td>Meaning System</td><td></td></tr>
<tr><td>Health Care System (Societal elements other than health health service system</td><td></td></tr>
<tr><td colspan="2">Relationship between elements</td><td></td></tr>
<tr style="display:none"></tr>
<tr><td rowspan="7">Methodological Approaches</td><td rowspan="6">Analytical Approaches</td><td>Historical analysis</td><td></td></tr>
<tr><td>Political Economy approach</td><td></td></tr>
<tr><td>Analysis of Power and Hierarchy</td><td></td></tr>
<tr><td>Epidemiological Orientation</td><td></td></tr>
<tr><td>Knowledge Pluralism</td><td></td></tr>
<tr><td>Health System Vantage Point</td><td></td></tr>
<tr><td colspan="2">Operational Approaches</td><td></td></tr>
<tr><td>Recommendations</td><td colspan="2"></td><td></td></tr>
<tr><td>Strengths</td><td colspan="2"></td><td></td></tr>
<tr><td>Limitations</td><td colspan="2"></td><td></td></tr>
<tr><td>Research Paradigm</td><td colspan="2"></td><td></td></tr>
</table>

2. Descriptive details of the selected studies

| |
|---|
| **Name of the study** |
| **Author/s** |
| **Year of Publication** |
| **Location (study site, if any)** |
| **Funding** |
| **Type of Research** |
| **Methodology of developing the framework** |
| **Structure of Disciplinary Interaction in the Study (Multi/Trans/Interdisciplinary)** |
| **Aim and Purpose of the study** |
| **Level of Analysis(Global/National/State/District)** |
| **Quality Of the Paper - Category (High/Medium/Low)** |

**The Health Systems Research Framework Version-1**

The Health Systems Research Framework (HSRF-1) prepared by reworking version-0 after the systematic narrative review of health systems frameoworks had two components: A descriptive component and an analytical component.

1. Descriptive component

| |
|---|
| **Name of the Study** |
| **Year of publication** |
| **Author/s** |
| **Institutions** |
| **Funding** |
| **Type of Institution** |

| |
|---|
| **Type of Research (Implementation/Evaluation/Perspective/review/ others)** |
| **Methodology of the study** |
| **Objective of the study** |
| **Level of analysis (Global/Inter-state/National/State/District)** |
| **Structure of Disciplinary Interaction (Multi/Trans/ Interdisciplinary)** |

2. Analytical component

| | | |
|---|---|---|
| | **Name of the study** | |
| | **Name of the conceptual framework** | |
| **Conceptualisation of health systems** | **Boundary** | |
| | **Sub-systems** | |
| | **Elements of health systems and its context** | **Ecosystems** |
| | | **Socio-political contexts (of health services)** |
| | | **Meaning systems** |
| | | **Health care system (societal elements other than health service system)** |
| | **Relationship between elements** | |
| **Methodological Approaches** | **Analytical Approaches** | **Historical Analysis** |
| | | **Political Economy of Health Services** |
| | | **Analysis of social stratification/power/hierarchy** |
| | | **Epidemiological orientation** |
| | | **Knowledge pluralism & Politics of knowledge** |

| | | |
|---|---|---|
| | | **Health System Vantage Point** |
| | **Operational Approaches** | |
| **Values and Principles** | **Effectiveness (Efficacy + coverage)** | |
| | **Technical Efficiency (Cost + time)** | |
| | **Allocative Efficiency** | |
| | **Sustainability** | |
| | **Equity** | |
| | **Autonomy** | |
| | **Safety** | |
| | **Clinical Rationality** | |
| | **Ethical Practice** | |
| | **Addressing Social Determinants of Health (SDH)** | |
| | **Flexibility/Decentralization** | |
| | **Others** | |
| **Outcomes** | | |
| **Goals** | | |
| **Recommendations** | | |
| **Research Paradigms** | | |
| **Emerging Issues** | | |

**The Health Systems Research Framework Version-2**

The Health Systems Research Framework (HSRF-2) prepared through the experience of applying HSRF version-1 to review studies from selected studies of various themes under HSR conducted in the Indian context, had three components: A Descriptive component, an Analytical component and a Research Coherence component.

1. Descriptive component

| |
|---|
| **Name of the Study** |
| **Year of publication** |
| **Author/s** |
| **Institutions** |
| **Funding** |
| **Type of Institution** |
| **Type of Research (Implementation/Evaluation/Perspective/review/ others)** |
| **Methodology of the study** |
| **Objective of the study** |
| **Level of analysis (Global/Inter-state/National/State/District)** |
| **Structure of Disciplinary Interaction (Multi/Trans/ Interdisciplinary)** |

2. Analytical component

| | | |
|---|---|---|
| | **Name of the study** | |
| | **Name of the conceptual framework** | |
| **Conceptualisation of health systems** | **Boundary** | |
| | **Sub-systems** | |
| | **Elements of health systems and its context** | **Ecosystems** |
| | | **Socio-political contexts (of health services)** |
| | | **Meaning systems** |
| | | **Health care system (societal elements other than health service system)** |
| | **Relationship between elements** | |
| **Methodological** | **Analytical** | **Historical Analysis** |

| | | |
|---|---|---|
| **Approaches** | **Approaches** | **Political Economy of Health Services** |
| | | **Analysis of social stratification/power/hierarchy** |
| | | **Epidemiological orientation** |
| | | **Knowledge pluralism & Politics of knowledge** |
| | | **Health System Vantage Point** |
| | **Operational Approaches** | |
| **Values and Principles** | **Effectiveness (Efficacy + coverage)** | |
| | **Technical Efficiency (Cost + time)** | |
| | **Allocative Efficiency** | |
| | **Sustainability** | |
| | **Equity** | |
| | **Autonomy** | |
| | **Safety** | |
| | **Clinical Rationality** | |
| | **Ethical Practice** | |
| | **Addressing Social Determinants of Health (SDH)** | |
| | **Flexibility/Decentralization** | |
| | **Others** | |
| **Outcomes** | | |
| **Goals** | | |
| **Recommendations** | | |

| |
|---|
| **Research Paradigms** |
| **Emerging Issues** |

3. Research coherence component

| |
|---|
| **Research Objective(s)** |
| **Research Question(s)** |
| **Values and Principles** |
| **Health Systems Problem Conceptualisation** |
| **Methodological Approach** |
| **Findings** |
| **Recommendations** |
| **Research Coherence** |
| **Strengths of Research** |

**The Dialogic Health Systems Research Framework (Health Systems Research Framework Version-3)**

The Dialogic Health Systems Research Framework has two components:

A. An initial description of the research, and

B. An assessment of research coherence

#### A. Description of the research

| |
|---|
| **Name of study/Title of report/paper** |
| **Year of publication** |
| **Author/s** |
| **Institutions** |
| **Funding** |
| **Type of Institution** |

| |
|---|
| **Type of Research / Purpose (Implementation/Evaluation/Perspective/ Review/ Policy Analysis/ Synthesising Research)** |
| **Objective of the study** |
| **Methodology of the study** |
| **Level of analysis (Global/Inter-state/National/State/District)** |
| **Area of analysis (Urban/rural/peri-urban/All)** |
| **Policy implications** |

**B. Assessment of Research Coherence (synthesised from analysis by three tools)**

| **Health Systems Research Components** | | **Research Coherence** | **Strengths of research** |
|---|---|---|---|
| Research Objective(s) | | | |
| Research Question(s) | | | |
| Values & principles for the Health System | | | |
| Health System Research conceptualisation | | | |
| Methodological approach | | | |
| Findings | | | |
| Recommendations | | | |

***Three Tools for Assessment of Research Coherence***

The assessment of research coherence is done through three background analytical steps:

1. Analysis of values and principles for the health system
2. Analysis of health system conceptualisation
3. Analysis of methodological approaches

The tools prepared for these three steps are given below:

1. **Tool for analysing values & principles**

| | **Values & Principles** | **Values & Principles Relevant to the Research**<br><br>**Yes / No**<br><br>**(a)** | **If Relevant : Present/ Limited/ Absent**<br><br>**(b)** | **Observations** (on how the incorporated values have been used in the research)<br>**(c)** |
|---|---|---|---|---|
| **Health System goals** | Sustainability | | | |
| | Equity | | | |
| | Context appropriateness | | | |
| | People-centredness | | | |
| **Health System functioning** | Effectiveness | | | |
| | Safe, Rational, Ethical Practice | | | |
| | Appropriate Technology | | | |
| | Technical efficiency | | | |
| | Affordability | | | |
| | Ecological sensitivity | | | |
| | Dignity in care | | | |
| | Self-reliance & Autonomy | | | |
| | Empowerment | | | |
| | Trust & transparency | | | |
| | Accountability & Responsiveness | | | |
| | Decentralisation (dialogic, deliberative, democratic) | | | |
| | Others | | | |

2. **Tool for analysing Health System Conceptualisation**

| | | | | | |
|---|---|---|---|---|---|
| **Conceptual framework**<br>**(name or briefly describe the framework)** | | | | | |
| | | | **Components Relevant to the Research (note those from options given in the previous column)**<br><br>**Yes/No**<br>**(a)** | **If Relevant: Present/Limited/Absent**<br><br>**(b)** | **Observations**<br><br>**(c)** |
| **Health System Conceptualisation** | **Boundary**<br>● Addressing Ecological & Social Determinants of Health<br>● Formal Health Service System<br>● Informal Health Care | | | | |
| | **Subsystems of formal health service system (Structures and processes)**<br>● Arrangements to address Ecological and Social Determinants<br>● Service delivery<br>● Health Workforce<br>● Information/knowledge<br>● Health technologies<br>● Finance<br>● Leadership & Governance<br>● Community engagement<br>● Others | | | | |
| | **Informal Health Services (Structures and processes)**<br>● Informal providers of allopathy<br>● Traditional caregivers of codified traditional systems<br>● Traditional caregivers of non-codified systems (Local Health Traditions) | | | | |
| | **Health System Vantage Point**<br>● Top-down/Institutional<br>● Bottom-up/Community centred<br>● Combined | | | | |
| | **Dynamic Elements of** | Ecosystems<br>● Interrelationships | | | |

| | | |
|---|---|---|
| | **Health System and its context** | between living organisms and their surroundings influencing health |
| | | Socio-political contexts of health and health care<br>● Combined social and political factors that influence people's health<br>● Socio-political factors that influence health care |
| | | Meaning systems of health and health care<br>● Collective and individual health-related worldviews, experiences and perceptions |
| | | Informal social arrangements & Community practices for health<br>● Health seeking behaviour<br>● Self-care<br>● Household-level care<br>● Societal arrangements (e.g. for leisure, exercise, sports)<br>● Community practices (e.g. seasonal foods; maternal & childcare)<br>● Emergent properties<br>● Others |
| | **Relationship of the elements across the system** | |
| | **Theory of change** | |

### 3. Tool for analysing Methodological Approaches

<table>
<tr><th colspan="2">Methodological Approaches</th><th>Approaches Relevant to the Research (note the relevant dimensions)<br>(a)</th><th>If Relevant: Present/Limited/Absent<br>(b)</th><th>Observations<br>(c)</th></tr>
<tr><td rowspan="5">Analytical approaches</td><td>Epidemiological approach (Health profile and determinants)</td><td></td><td></td><td></td></tr>
<tr><td>Historical analysis</td><td></td><td></td><td></td></tr>
<tr><td>Social Science approaches<ul><li>Political Science approach (e.g. Political economy of health)</li><li>Sociological approach</li><li>Social geographic approach</li><li>Economic analysis of health care</li><li>Social Anthropological approach</li><li>Knowledge pluralism and politics of knowledge</li><li>Analysis of social stratification/power/hierarchy</li><li>Others</li></ul></td><td></td><td></td><td></td></tr>
<tr><td>Management Science approaches</td><td></td><td></td><td></td></tr>
<tr><td>Ecosystem approaches</td><td></td><td></td><td></td></tr>
</table>

| |
|---|
| **Operational approaches***<br>● Intervention study design<br>● Non-Intervention study design |
| **Nature of disciplinary interaction**<br>● Unidisciplinary<br>● Multidisciplinary<br>● Interdisciplinary<br>● Transdisciplinary |
| **Ethical Considerations of the Study**<br>**(Consent, Confidentiality, Conflict of Interest)** |
| **Quality Assessment of Research Operationalisation**** |
| **Research paradigm**<br>● Conventional Public Health/Pragmatist<br>● Positivist<br>● Realist closer to Positivist<br>● Realist<br>● Realist closer to Holist<br>● Holist |

Note:

* Operational approaches will have to be further detailed depending on the methodology applied

** Any quality assessment tool deemed relevant for the research can be used
