## Supplementary 2: Definition of HSR for "The Dialogic Health Systems Research Framework (DHSRF): A tool for facilitating self-criticality, researcher interactions and knowledge management in Health Systems Research & Policy Studies"

The working paper for WHO Global Strategy on Health Systems Research by Hoffman et al. (2012) defines HSR as "a multidisciplinary field of health research which studies governance, financial and delivery arrangements for health care and public health services, implementation considerations for reforming or strengthening these arrangements, and broader economic, legal, political and social contexts in which these arrangements are negotiated and operate. Health systems research aims to improve the understanding and performance of health systems. Health systems research includes all of health services research, most health policy research, and some clinical and population health research, but does not include any biomedical research." It sets out the need to simultaneously address micro, meso and macro level dimensions.

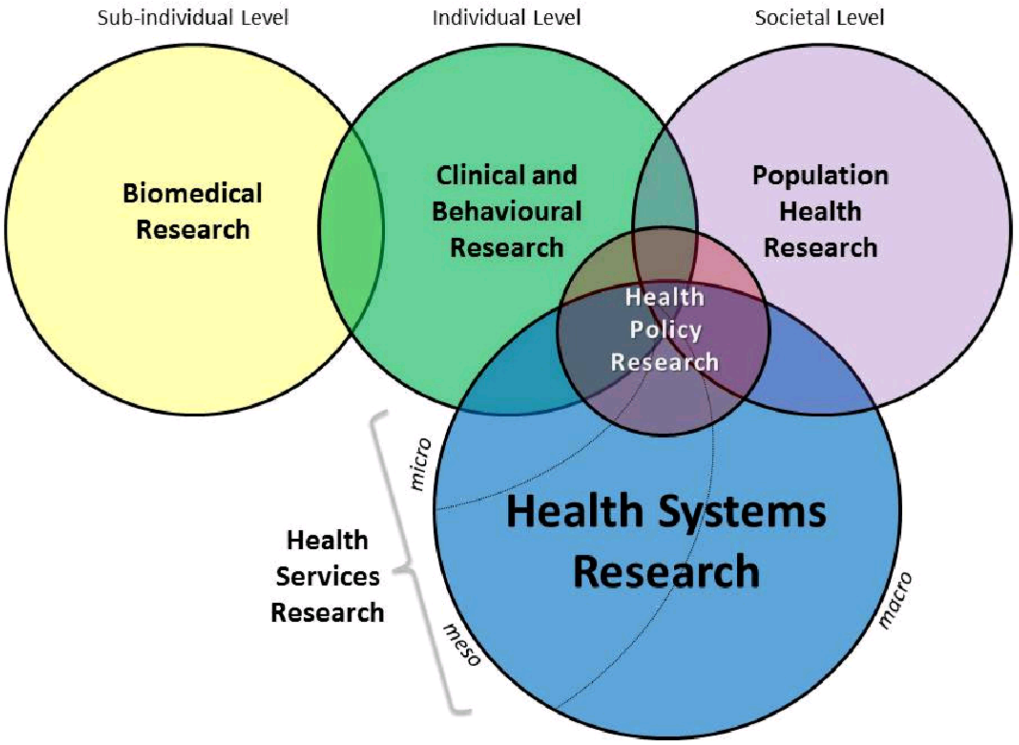


Figure 1. Health Systems Research as a Multidisciplinary Field of Health Research (Source: Hoffman et al., 2012)

The Alliance for Health Policy and Systems Research (AHPSR), gaining prominence in the decade of the 2010s define their proposed iteration of HSR, Health Policy and Systems Research (the nomenclature of the field adopted by WHO as a member of this alliance), as a field " … that seeks to understand and improve how societies organize themselves in achieving collective health goals, and how different actors interact in the policy and implementation processes to contribute to policy outcomes. By nature, it is interdisciplinary, a blend of economics, sociology, anthropology, political science, public health and epidemiology that together draw a comprehensive picture of how health systems respond and adapt to health policies, and how health policies can shape – and be shaped by – health systems and the broader determinants of health. "

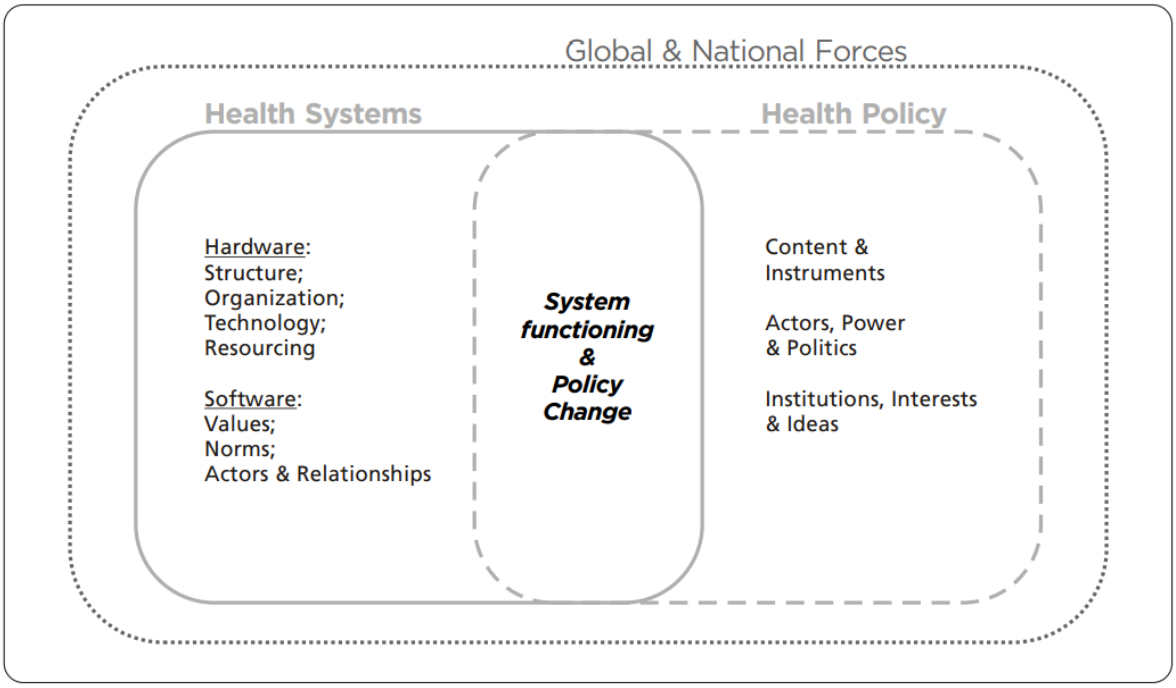


Figure 2. The terrain of Health Policy and Systems Research (Source: Gilson, 2012)

While we broadly agree with Hoffman et al's definition, including that bio-medical research is outside the remit of HSR and Policy Studies, we believe that examining 'the system of biomedical research' needs to be explicitly included within HSR. Epidemiology, the cornerstone of public health, includes biomedical research besides other dimensions. Both biomedical and clinical research directly influence people's health through health technologies and other interventions that are delivered through health systems. The biomedical research system thus should be considered an important component of HSR.

On the other hand, we differ from the definition of HPSR which puts Health Policy as the central objective of HSR. Health Policy is a crucial part of HSR but that is not the only imperative of HSR. Policy studies (PS) is one of the many components of HSR, with implementation research and the socio-cultural dimensions of health care as other major components. In our view, HSR and PS are two closely related and often overlapping, yet distinct interdisciplinary fields with widely differing methodological imperatives. While HSR is mainly concerned with the technical and operational dimensions of planning and implementation, Health Policy Studies are more oriented towards the political dimensions and processes of policy making about how to improve population health and the various systems/sub-systems that may be involved. Therefore, we prefer to keep them linked and yet stand apart as HSR & PS. From a Public Health perspective, epidemiology, HSR and policy studies are an intrinsic part of the discipline and the linkages between them have to be considered even when any one of them is the focus of study. The socio-cultural dimensions form the context in which the institutional systems perform, and which they influence. This tends to be the least examined in HSR and PS despite it being recognised as a significant barrier and facilitator of policy and implementation. Here we use HSR as a composite of all three, even while we prefer to make the distinction by expanding it to HSR & PS.

### References to Supplementary 2

Hoffman, S. J., Røttingen, J. A., Bennett, S., Lavis, J. N., Edge, J. S., & Frenk, J. (2012). Background paper on conceptual issues related to health systems research to inform a WHO global strategy on health systems research. *Health Systems Alliance*.https://www.academia.edu/65618679/Background_Paper_on_Conceptual_Issues_Related_to_Health_Systems_Research_to_Inform_a_WHO_Global_Strategy_on_Health_Systems_Research

Gilson, L., & World Health Organization. (2013). *Health policy and system research: a methodology reader: the abridged version*. World Health Organization. https://iris.who.int/handle/10665/44803
