## Supplementary 3: Glossary for "The Dialogic Health Systems Research Framework (DHSRF): A tool for facilitating self-criticality, researcher interactions and knowledge management in Health Systems Research & Policy Studies"

The glossary contains standard definitions of most terms as found in the literature. In some cases they have been modified by the research team based on review and analysis to appropriately reflect their positions. Those using the DHSRF can use these definitions to create their own operational definitions as relevant to their research.

**Health Systems Research -** Health systems research is a multi-disciplinary field of health research which studies governance, financial and delivery arrangements for health care and public health services, implementation considerations for reforming or strengthening these arrangements, and broader economic, legal, political and social contexts in which these arrangements are negotiated and operated. The purpose of health systems research is to improve the understanding and performance of health systems. Health systems research includes all of health services research, most health policy research, and some clinical and population health research, but does not include any biomedical research.[1]

**Health Policy and Systems Research -** Health policy and systems research (HPSR) is an emerging field that seeks to understand and improve how societies organize themselves in achieving collective health goals, and how different actors interact in the policy and implementation processes to contribute to policy outcomes. By nature, it is inter-disciplinary, a blend of economics, sociology, anthropology, political science, public health and epidemiology that together draw a comprehensive picture of how health systems respond and adapt to health policies, and how health policies can

[1] Hoffman, J.S., Rottingen, J., Bennett, S., Lavis, J.N., Edge, J.S. & Frenk, J. (2012). Background Paper on Conceptual Issues Related to Health Systems Research to Inform a WHO Global Strategy on Health Systems Research. https://d1wqtxts1xzle7.cloudfront.net/77136817/alliancehpsr_hsrstrategy_conceptualpaper-libre.pdf?1640293274=&response-content-disposition=inline%3B+filename%3DBackground_Paper_on_Conceptual_Issues_Re.pdf&Expires=1708932463&Signature=f-8Z6jyjesQKmDb6Zvz9rspcKllhZyMzJPFcAsFbsjj98QU2QhI8JO~z3aqtK0HYPVKBOmyAjPnXjp1z85CoosaYyBJd2-N1clFIvTQztirxMyxc73pC3nDf~7U5KLzG9sziwFuo5Crfp3j6V~CXu3kNzePTQCU5WL-TKvs~rbyLbuIEKiJ7MUcugtcJnGhGE79JqEAJokm5z0JZD~7qcNXxEcSefbmCKur9GahaFMcCH2rPytTjPjnQdwFFY0M7Fo~FC4Kk-6ys2mYBCMaj6Ky5KyjZSIwxi1mxGn9pNEIQDjr0~gxy0342BCrVoQ9OhoZ9ahq8GDOneLn~AVqSug__&Key-Pair-Id=APKAJLOHF5GGSLRBV4ZA)

shape – and be shaped by – health systems and the broader determinants of health.[2]

**Health Systems Research and Policy Studies** - HSR and PS are two closely related and yet distinct interdisciplinary fields. HSR is more oriented towards the technical and operational dimensions of planning and implementation while Health Policy Studies are more oriented towards the political dimensions and processes of policy decision making about how to improve population health and the various systems and subsystems that may be involved. Therefore, we prefer to keep them linked and yet stand apart as HSR and PS. From a Public Health perspective, epidemiology, HSR and policy studies are an intrinsic part of the discipline and the linkages between them have to be considered even when any one of them is the focus of study.[3]

1. **Frameworks**

A. **Health System Frameworks** - A bird's eye view of the health system that defines, describes and explains the health system, its objectives, structural and organizational elements, functions and processes.[4]

B. **Health System Research Framework** - A framework that elucidates the major components of health systems research (research objective, Health systems conceptualisation, methodological approach, data analysis and findings, recommendations, and values and principles underlying all these) and examines their coherence across the research.

[2] Alliance for Health Policy and Systems Research, https://ahpsr.who.int/what-we-do/what-is-health-policy-and-systems-research-(hpsr)

[3] Dever, G.E.A. An epidemiological model for health policy analysis, 1976, Soc Indic Res 2, 453–466. https://doi.org/10.1007/BF00303847

[4] Shakarishvili, G., Atun, R., Berman, P., et.al Converging Health Systems Frameworks: Towards A Concepts-to-Actions Roadmap for Health Systems Strengthening in Low and Middle Income Countries. Global Health Governance, 2010, Vol. III, No. 2, http://blogs.shu.edu/wp-content/blogs.dir/109/files/2011/11/Shakarishvili-et-al_Converging-Health-Systems-Frameworks_Spring-2010.pdf

C. **Research Coherence:** Research Coherence "describes the fit between the aim, the philosophical perspective adopted, and the researcher role in the study as well as the methods of investigation, analysis and evaluation undertaken by the researcher."[5]

2. **Health System Approaches**

A. **Techno-managerial approach**: An approach to health systems that limits its focus to the organisation of technology-based health care and management of formal delivery systems, with minimal attention to social dimensions and processes shaping health and health care.[6]

B. **Comprehensive/Socio-cultural approach:** An approach to health systems that addresses the different societal arrangements that exist for maintaining and improving people's health, including the formal and informal arrangements. These include macro, meso and micro level arrangements related to food, hygiene, sanitation, physical exercise, leisure and social relationships, expression of emotions, etc., in addition to the specific practices for prevention of disease and for promoting health, treating ill-health and easing physical and mental suffering, thereby going well beyond consideration of the formally organised health services. It includes the study of the formal health services as social institutions embedded in this socio-cultural context.[7]

3. **Health system definition**:

A. **Techno-managerial definition:** A health system consists of all organisations, people and actions whose primary intent is to promote, restore or maintain health.[8]

B. **Socio-cultural definition:** The systemic determinants of health in a population that generate its health and morbidity profile as well as the health care systems developed to maintain health and deal with ill health.

---

[5] Vaismoradi, M., & Salsal, D. mahvash. (2011). Coherence in qualitative research. *Advances in Nursing & Midwifery*, *20*(70), 45–53. Retrieved from https://journals.sbmu.ac.ir/en-jnm/article/view/2047

[6] Luning, P.A. & Marcelis, W.J. (2009). A food management research methodology integrating technological and managerial theories. Trends in Food Science & Technology, 20(1), 35-44. https://doi.org/10.1016/j.tifs.2008.09.013

[7] Purola, T. (1972). A Systems Approach to Health and Health Policy. Medical Care, 10(5), 373–379. http://www.jstor.org/stable/3763004

[8] World Health Organisation, https://www.who.int/india/health-topics/health-systems-governance

Adopting the Socio-cultural definition:

A. **Health care system**: The health care system is a whole of political, economic and cultural, technical and organizational factors, relations, processes and elements, in which individuals, groups and communities interrelate, having the goal to satisfy their health needs.[9]

B. **Health service system**: Health services are any service (i.e. not limited to medical or clinical services) aimed at contributing to improved health or to the diagnosis, treatment and rehabilitation of sick people.[10]

C. **Medical care system**: The component of the health service system that is organised around the delivery of *medical* services.

D. **Boundary of the Health System:** The outer limits (context, institutions, capacities) within which the health system operates.[11]

E. **Sub-Systems of formal Health Service System:** This constitutes both structures and processes across components of the Health Service System. From a techno-managerial perspective, these include the Formal aspects of the ecological and social determinants of health, Leadership and governance; Service delivery; Health workforce; Information/knowledge; Health Technologies; Finance[12] and Community engagement.

Various subsystems of the health care system viewed from a socio-cultural perspective include the following[13]:

---

[9] Kovačić, L. & Jakšić, Z. "Health Care as a System: Elements, Boundaries, Levels", in Kovačić, L & Zaletel-Kragelj, L. (Ed.) Management in Health Care Practice: A Handbook for Teachers, Researchers and Health Professionals, 2008, Hans Jacob Publishing Company: Zagreb, https://www.biejournals.de/index.php/seejph/article/download/1910/pdf_4/6732

[10] World Health Organisation, Health Systems Strengthening Glossary, https://www.who.int/docs/default-source/documents/health-systems-strengthening-glossary.pdf

[11] WHO, Health Systems Strengthening Glossary, https://www.who.int/docs/default-source/documents/health-systems-strengthening-glossary.pdf

[12] WHO Health System Building Blocks, https://extranet.who.int/nhptool/BuildingBlock.aspx

[13] Kovačić, L. & Jakšić, Z. "Health Care as a System: Elements, Boundaries, Levels", in Kovačić, L & Zaletel-Kragelj, L. (Ed.) Management in Health Care Practice: A Handbook for Teachers, Researchers and

- Socio-political subsystem - the main health legislation is as a rule at the national level, but communities could be more or less self-reliant and responsible for planning and organization of health care. Solidarity and support is usually at higher levels;
- Subsystem of users (communities and individuals) - responsibility and participation of the community in planning, organization, operation and control;
- Socioeconomic subsystem - health insurance (obligatory, voluntary, private), and private relation of health providers and users;
- Managerial subsystem (decision making process): level of autonomy of health institutions, type of management (autocratic, birocratic, corporative laissez-faire);
- Technological subsystem - Comprehensive approach in provision of primary health care, segmented at secondary level;
- Organizational subsystem – levels of the health infrastructure (primary, secondary, tertiary), type of health institutions (individual practices, group practices, health centres, day hospitals, clinical hospitals);
- Health care infrastructure (health care facilities) - infrastructure could be a subsystem which supports the operations of an organization (health centre, health sub-centre, hospital, medical centre, institute of public health, rehabilitation centre and spas, pharmacy, specialized institutes - vaccine production, emergency services in large cities, blood supply, etc, private practice - dentists, physicians, nurses, herbalists and other alternative practitioners);
- Supporting systems - training and research institutes, health related industries (production of drugs, equipment, etc.).

### 4. Health Systems elements and analytical approaches

#### Dynamic Elements of a Health System and Its context

A. **Ecosystem** - The dynamic complex of living organisms, their surroundings, and all their interrelationships contributing to people's health or the lack thereof.[14]

---

Health Professionals, 2008, Hans Jacob Publishing Company: Zagreb, https://www.biejournals.de/index.php/seejph/article/download/1910/pdf_4/6732

[14] Britannica, The Editors of Encyclopaedia. "ecosystem". Encyclopedia Britannica, 12 Feb. 2024, https://www.britannica.com/science/ecosystem. Accessed 26 February 2024

B. **Socio-political context** - The combined social and political factors that influence people's health and well-being by shaping the conditions in which people live and work as well as their access to essential social and economic resources.

C. **Meaning systems** - The collective and individual health-related worldviews, experiences and perceptions about health, health problems and health care.

- **Health Culture** - Health culture covers a wide range of considerations which intimately interact with one another to form a sub-cultural complex. Cultural perceptions of health problems, their cultural meanings and the cultural response to these problems, both in terms of formation of various institutions to deal with various health problems and actual (health) behaviour of individuals and groups form this sub-cultural complex.[15]

D. **Informal social arrangements & Community practices for health** - All informal societal arrangements for maximising health and minimising ill-health, shaped by agencies of different actors, their interactions and emergent behaviours. This includes informal care, people's health preserving and improving practices, social support in ill-health or vulnerabilities.

**Emergent Behavior** - The spontaneous creation of order, which appears when smaller entities on their own jointly contribute to organized behaviours as a collective, resulting in the whole being greater and more complex than the sum of the parts.[16]

**Health system vantage point -** Analysis of the perspective underlying methods of health systems research, planning and policymaking, defining objectives and ways of achieving them. A health system vantage point can be either bottom-up or top-down, or a combination of the two.

---

[15] Banerji, D. (1985). Health and Family Planning Services in India: An Epidemiological, Socio-cultural, and Political Analysis and a Perspective. Lok Paksh, New Delhi.

[16] Paina, L. & Peters, D.H. (2012). Understanding pathways for scaling up health services through the lens of complex adaptive systems. Health Policy and Planning, 27(5), 365-73, https://pubmed.ncbi.nlm.nih.gov/21821667/

- **Bottom-up vantage point** - A bottom-up or social vantage point encourages research, evaluation and planning beginning at the grassroots (community level)—with the people who are meant to be the primary beneficiaries of the health system with all the components that prevail in their perception and practice— with reference to progressive levels of the Health System. It grants them rationality and agency and attempts to understand these.

- **Top-down vantage point** - A top-down/institutional vantage point, on the other hand, starts at the top (beginning at the institutional level) —with the planners and policymakers and what shapes their decisions —followed by dissemination of decisions and implementation down the rest of the levels up to the community. It gives primacy to formal institutions and activities and their functional attributes.

- **A Combination** - Both Bottom-up and Top-down approaches are applied, holistically bridging the two.

**Theory of Change -** Theory of Change is "an outcomes-based approach which applies critical thinking to the design, implementation, and evaluation of initiatives and programs intended to support change in their context".[17]

**Analytical approaches**

A. **Historical analysis -** Historical analysis involves analyzing the different cause-and-effect relationships present in each scenario, considering the ways individuals, influential ideas, and different mindsets interact and affect one another.[18]

---

[17] Vogel I. Review of the Use of "Theory of Change" in International Development. London: Commissioned by the UK Department for International Development; 2012. As cited in, Paina, L., Wilkinson, A., Tetui, M. et al. (2017). Using Theories of Change to inform implementation of health systems research and innovation: experiences of Future Health Systems consortium partners in Bangladesh, India and Uganda. Health Research Policy and Systems, 15 (suppl 2), 109. https://doi.org/10.1186/s12961-017-0272-y

[18] Cole, S., Breuer, K., Palmer, S.W. & Blakeslee, B. "How History is Made: A Student's Guide to Reading, Writing, and Thinking in the Discipline", September 2022, Mavs Open Press, University of Texas at Arlington Libraries,
https://uta.pressbooks.pub/historicalresearch/chapter/what-is-historical-analysis/#:~:text=The%20very%20essence%20of%20historical,interact%20and%20affect%20one%20another.

B. **Political Economy of Health -** The "political economy of health" is concerned with how political and economic domains interact and shape individual and population health outcomes.[19]

C. **Analysis of social stratification, power & hierarchy** - Analysis of social inequalities and its impact on health and health care.

D. **Epidemiological orientation** - Consideration of epidemiological context of the health system being studied.

E. **Knowledge pluralism -** The existence of different forms of knowledge related to individual and collective health, including the bio-medical and social sciences, the several traditions of knowledge for understanding, improving and maintaining health such as Biomedicine, Ayurveda, Unani, Siddha, Traditional Chinese Medicine, Chiropractice, traditional birth attendants and home remedies.

- **Medical pluralism -** Medical pluralism describes the availability of different medical approaches, treatments, and institutions that people can use while pursuing health: for example, combining biomedicine with so-called traditional medicine or alternative medicine.[20]

- **Politics of Knowledge -** The privileging of certain forms of knowledge, and consequent de-legitimizing of other forms – under particular discourses and the resultant institutional arrangements is referred to as the Politics of knowledge.[21]

### 5. Values and Principles for Health Systems

- **Sustainability (Financial sustainability of program/economic viability,**

[19] Harvey, M. (2021). The Political Economy of Health: Revisiting Its Marxian Origins to Address 21st-Century Health Inequalities, American Journal of Public Health, 111(2), 293-300. https://doi.org/10.2105/AJPH.2020.305996
[20] Khalikova, V. (2021) 2023. "Medical pluralism". In The Open Encyclopedia of Anthropology, edited by Felix Stein. Facsimile of the first edition in The Cambridge Encyclopedia of Anthropology. Online: http://doi.org/10.29164/21medplural
[21] Gaitonde R, et.al (2019) Some Thoughts on Health for All: the rationale for engaging with the politics of knowledge. MFC Bulletin, 380:9–15, https://www.mfcindia.org/mfcpdfs/MFC380.pdf

**environmental integrity and social justice/equity/health outcome) -** The potential for sustaining beneficial outcomes for an agreed period at an acceptable level of resource commitment within acceptable organizational and community contingencies.[22] A sustainable health system improves population health by continually delivering the key functions of providing services, generating resources, financing and stewardship, incorporating principles of financial fairness, equity in access, responsiveness and efficiency of care, and does so in an environmentally sustainable manner.[23]

- **Equity -** (i) the absence of systematic or potentially remediable differences in health status, access to healthcare and health‑enhancing environments, and treatment in one or more aspects of health across populations or population groups defined socially, economically, demographically or geographically within and across countries. (ii) a measure of the degree to which health policies are able to distribute well‑being fairly.[24]

- **Context-Appropriateness** - Context is conceptualized as a set of characteristics and circumstances that consist of active and unique factors that surround the implementation. As such it is not a backdrop for implementation but interacts, influences, modifies and facilitates or constrains the intervention and its implementation. Context is usually considered in relation to an intervention or object, with which it actively interacts. A boundary between the concepts of context and setting is discernible: setting refers to the physical, specific location in which the intervention is put into practice. Context is much more versatile, embracing not only the setting but also roles, interactions and relationships.[25]

---

[22] WHO, Health Systems Strengthening Glossary, https://www.who.int/docs/default-source/documents/health-systems-strengthening-glossary.pdf
[23] Gocke, D., Johnston-Webber, C., McGuire, A., & Wharton, G. "Building Sustainable and Resilient Health Systems: Key Findings from Country Reports", Partnership for Health System Sustainability and Resilience (PHSSR). May 2023, https://www3.weforum.org/docs/WEF_PHSSR_Building_Sustainable_and_Resilient_Health_Systems_2023.pdf )
[24] WHO, Health Systems Strengthening Glossary, https://www.who.int/docs/default-source/documents/health-systems-strengthening-glossary.pdf
[25] Pfadenhauer, L., Rohwer, A., Burns, J., Booth, A., Lysdahl, K.B., Hofmann, B., et al. (2016) Guidance for the assessment of context and implementation in health technology assessments (HTA) and systematic reviews of complex interventions: the Context and Implementation of Complex Interventions (CICI) framework. INTEGRATE-HTA Consortium, http://www .integrate-hta.eu/downloads/

- **People-centred care/People-centredness** - Care that is focused and organized around the health needs and expectations of people and communities rather than on diseases. People‑centred care extends the concept of patient‑centred care to individuals, families, communities and society. Whereas patient‑centred care is commonly understood as focusing on the individual seeking care—the patient, people‑centred care encompasses these clinical encounters and also includes attention to the health of people in their communities and their crucial role in shaping health policy and health services.[26]

- **Effectiveness -** Effectiveness is the extent to which a specific intervention, procedure, regimen or service, when deployed in the field in routine circumstances, does what it is intended to do for a specified population.[27]

- **Safety -** Within the broader health system context, it is a framework of organized activities that creates cultures, processes, procedures, behaviours, technologies and environments in health care that consistently and sustainably lower risks, reduce the occurrence of avoidable harm, make error less likely and reduce impact of harm when it does occur.[28]

- **Clinical rationality -** Rationality is commonly defined as decision making that helps us achieve our goals. In the context of clinical medicine, this typically means the desire to improve our health. Rationality does not guarantee that a decision is error free; rather, rational decision-making accounts for the potential consequences of possible errors of action—false negatives and false positives—to help arrive at optimal outcomes.[29]

- **Ethical Practice -** This is concerned with the obligations and practices of the health

---

[26] WHO, Health Systems Strengthening Glossary, https://www.who.int/docs/default-source/documents/health-systems-strengthening-glossary.pdf
[27] WHO, Health Systems Strengthening Glossary, https://www.who.int/docs/default-source/documents/health-systems-strengthening-glossary.pdf
[28] World Health Organisation, https://www.who.int/news-room/fact-sheets/detail/patient-safety
[29] Djulbegovic, B., Elqayam, S. & Dale, W. (2018). Rational decision making in medicine: Implications for overuse and underuse. Journal of Evaluation in Clinical Practice, 24(3), 655-665. doi: 10.1111/jep.12851

care professionals and institutions to the patient and society.[30]

- **Appropriate Technology -** Appropriate technology is defined as the adaptation to local circumstances and conditions of knowledge and skills which are scientifically sound and acceptable to those who apply them and those for whom they are used. It should be affordable and should include appropriate use and effective interaction between service users and performers, as well as control of the cost and clinical benefits. Appropriate technology does not mean primitive or necessarily simple and/or less expensive. The initial cost should be considered within the context of the overall benefits and the expected outcome in the long run. Priority should be given to technologies improving public health services, with emphasis on equal access to health care for all.[31]

- **Technical Efficiency -** Technical efficiency is concerned with achieving maximum outputs with the least cost (monetary and time), and thereby informs health systems design.[32]

- **Affordability -** A system for financing health services so people do not suffer financial hardship when using them.[33] Health facilities, goods and services must be affordable for all. Payment for health-care services, as well as services related to the underlying determinants of health, has to be based on the principle of equity, ensuring that these services, whether privately or publicly provided, are affordable for all, including socially disadvantaged groups. Equity demands that poorer households should not be disproportionately burdened with health expenses as compared to richer households.[34]

---

[30] Markose, A., Krishnan, R. & Ramesh, M. (2016). Medical ethics. Journal of Pharmacy and Bioallied Sciences, 8(Suppl 1), S1-S4. doi: 10.4103/0975-7406.191934. https://www.ncbi.nlm.nih.gov/pmc/articles/PMC5074007/

[31] World Health Organisation, https://applications.emro.who.int/docs/em_rc44_tech_disc_1_en.pdf

[32] Akazili, J., Adjuik, M., Chatio, S., Kanyomse, E., Hodgson, A., Aikins, M. & Gyapong, J. (2008) What are the Technical and Allocative Efficiencies of Public Health Centres in Ghana? Ghana Medical Journal, 42(4):149-55. https://www.ncbi.nlm.nih.gov/pmc/articles/PMC2673839/

[33] WHO, https://www.who.int/news-room/questions-and-answers/item/what-is-universal-health-coverage

[34] The measurement and monitoring of water supply, sanitation and hygiene (WASH) affordability: a missing element of monitoring of Sustainable Development Goal (SDG) Targets 6.1 and 6.2. New York: United Nations Children's Fund (UNICEF) and the World Health Organization, 2021, https://www.who.int/publications/i/item/9789240023284

- **Ecological Sensitivity -** Environmental sensitivity is related to an interest in the environment and presenting behaviours to protect it; it is a very important influence in ensuring sustainable development.[35]

- **Dignity in Care -** The right of individuals to be treated with respect as persons in their own right.[36]

- **Self-Reliance -** The capacity of individuals, communities or national authorities to take the initiative in assuming responsibility for their own health development and adopting adequate measures to maintain health that are understood by them and acceptable to them, knowing their own strengths and resources and how to use them and knowing when, and for what purpose, to turn to others for support and cooperation.[37]

- **Autonomy -** The right of patients to make decisions about their medical care without their health care provider trying to influence the decision. Patient autonomy does allow for health care providers to educate the patient but does not allow the health care provider to make the decision for the patient.[38]

- **Empowerment -** In health promotion, empowerment is a process through which people gain greater control over decisions and actions affecting their health.[39] Health empowerment is a cornerstone of a patient-centered approach to healthcare. Empowerment allows patients to take the initiative in making decisions about their own health care and quality of life, rather than passively complying with decisions made by others.[40]

---

[35] Yayla, O., Keskin, E. & Keles, H. (2022) "The Relationship Between Environmental Sensitivity, Ecological Attitude, and the Ecological Product purchasing Behaviour of Tourists" European Journal of Tourism, Hospitality and Recreation, 12(1), pp. 31-45. https://doi.org/10.2478/ejthr-2022-0002

[36] WHO Centre for Health Development (2004). A glossary of terms for community health care and services for older persons, https://apps.who.int/iris/handle/10665/68896

[37] WHO Centre for Health Development (2004). A glossary of terms for community health care and services for older persons, https://apps.who.int/iris/handle/10665/68896

[38] MedicineNet as cited in Harvard Health Publishing, https://www.health.harvard.edu/blog/take-control-of-your-health-care-exert-your-patient-autonomy-2018050713784

[39] WHO, Health Promotion Glossary of Terms 2021, https://www.who.int/publications/i/item/9789240038349

[40] Jiang, F., Liu, Y., Hu, J., & Chen, X. (2022) Understanding Health Empowerment From the Perspective of Information Processing: Questionnaire Study. Journal of Medical Internet Research 24(1):e27178. https://www.ncbi.nlm.nih.gov/pmc/articles/PMC8790685/

- **Trust -** Trust in health care is usually defined as a set of expectations that the patient has from the doctor and the healthcare system to help them heal. This set of expectations includes appropriate diagnosis, correct treatment, non-exploitation, genuine interest in the welfare of the patient and transparent disclosure of all information. Trust is like a forward-looking covenant between the doctor and the patient.[41]

- **Transparency -** The Institute of Medicine (IOM) defines healthcare transparency as making available to the public, in a reliable, and understandable manner, information on the health care system's quality, efficiency and consumer experience with care, which includes price and quality data, so as to influence the behavior of patients, providers, payers, and others to achieve better outcomes (quality and cost of care).[42]

- **Accountability** – The result of the process which ensures that health actors take responsibility of what they are obliged to do and are made answerable for their actions.[43]

- **Responsiveness -** Responsiveness entails reacting effectively to the needs and demands of the population and its different subpopulations and vulnerable groups. The content of the minimum package of activities should be informed both by the burden of disease and by the perceived needs of the population. It is a function of governance weighing the technical arguments; perceived needs; existing values and principles, and to decide which trade-offs to make, taking into account the infrastructure, level of development and capacity of implementation.[44]

---

[41] Gopichandran, V. & Chetlapalli, S.K. (2013). Dimensions and determinants of trust in health care in resource poor settings--a qualitative exploration. PLoS One, 8(7): e69170. https://www.ncbi.nlm.nih.gov/pmc/articles/PMC3712948/

[42] American College of Physicians. "Healthcare Transparency – Focus on Price and Clinical Performance Information", 2010. https://www.acponline.org/acp_policy/policies/healthcare_transparency_2010.pdf

[43] UHC 2030, Health budget literacy, advocacy and accountability for universal health coverage Toolkit for capacity-building, https://www.uhc2030.org/fileadmin/uploads/uhc2030/2_What_we_do/2.3_Sharing_knowledge_and_networks/2.3.3_Civil_society_engagement/Health_Budget_Literacy/WHO013_UHC2030-capacity-building-toolkit_glossary.pdf

[44] World Health Organization (2000). The World Health Report 2000. Improving Performance, World Health Organization, Geneva. https://www.who.int/publications/i/item/924156198X

- **Decentralization -** Political reform designed to promote local autonomy, decentralization entails changes in authority and financial responsibility for health services. Hence, decentralization can have a large impact on health service performance. There are several forms of decentralization affecting the health sector in different ways: (i) deconcentration, which transfers authority and responsibility from the central level of the Ministry of Health to its field offices; (ii) delegation, which transfers authority and responsibility from the central level of the Ministry of Health to organizations not directly under its control; (iii) devolution, which transfers authority and responsibility from the central level of the Ministry of Health to lower level autonomous units of government; (iv) privatization, which involves the transfer of ownership and government functions from public to private bodies, which may consist of voluntary organizations and for-profit and not-for-profit private organizations, with varying degree of government regulation.[45] Decentralization as an 'intervention' is often used in the health systems literature—as an arrangement in which the power, resources or responsibilities are transferred from central to peripheral actors.[46]

### 6. Health System Research Paradigms

**Paradigm -** A paradigm constitutes a set of theories, assumptions, and ideas that contribute to one's worldview and approach to engaging with other people or things. It is the lens through which a researcher views the world and examines the methodological components of their research to make a decision on the methods to use for data collection and analysis.[47]

**Research paradigm -** Practical application of the various elements of the paradigm in health systems research. Research paradigms consist of four philosophical elements: axiology, ontology, epistemology, and methodology.[48] These four elements inform the design and

---

45 WHO, Health Systems Strengthening Glossary, https://www.who.int/docs/default-source/documents/health-systems-strengthenWHing-glossary.pdf

46 Mills, A., Vaughan, J.P., Smith, D.L & Tabibzadeh, I. (1990). Health System Decentralization: Concepts, Issues and Country Experience. Geneva: World Health Organization

47 Kivunja, C. & Kuyini, A.B. (2017) Understanding and applying research paradigms in educational contexts. International Journal of Higher Education. 2017;6(5):26-41. As cited in: Alele, F., & Malau-Aduli, B. (2023). *An introduction to research methods for undergraduate health profession students.* James Cook University. https://jcu.pressbooks.pub/intro-res-methods-health

48 Creswell JW. Educational Research: Planning, Conducting, and Evaluating. 4th ed. W. Ross MacDonald School Resource Services Library; 2013

conduct of research projects (Figure 1.1), and a researcher would have to consider the paradigms within which they would situate their work before designing the research.[49]

A. **Conventional public health/Pragmatist -** A paradigm relying on framing the health problem from an epidemiological and problem-solving approach, designing levels of prevention to deal with it from structural to the individual level. Linkages with societal processes and context are minimally defined with little social theory being employed. Primarily quantitative methods combined with apriori social and organisational knowledge. Epistemologically, this paradigm follows a pragmatist stance that is focused on practical application of knowledge to real-world settings.[50]

B. **Positivist -** Positivism relies on the hypothetico-deductive method to verify a priori hypotheses that are often stated quantitatively, where functional relationships can be derived between causal and explanatory factors (independent variables) and outcomes (dependent variables).[51]

C. **Realist -** This theoretical paradigm is informed by a branch of philosophy called Critical Realism that distinguishes between the 'real' world and the 'observable' world, giving importance to **context** which triggers certain **mechanisms** in turn producing observable **outcomes**. Using plural methodology and an explanatory approach (what works? for whom? and why? Similarly, for what does not work as well) it explores mechanisms linked to actors and contexts mediating cause and effect.[52]

D. **Realist closer to Positivist** - Narrow boundary, context-mechanism-outcome with minimal attention to context and linkages; mixed methods with greater role of the quantitative; Top-down

---

[49] Alele, F., & Malau-Aduli, B. (2023). An introduction to research methods for undergraduate health profession students. James Cook University. https://jcu.pressbooks.pub/intro-res-methods-health)https://jcu.pressbooks.pub/intro-res-methods-health

[50] Creswell JW. Educational Research: Planning, Conducting, and Evaluating. 4th ed. W. Ross MacDonald School Resource Services Library; 2013

[51] Park, Y.S., Konge, L., Artino Jr, A.R. (2020). The Positivism Paradigm of Research. Academic Medicine 95(5), 690-694. https://journals.lww.com/academicmedicine/fulltext/2020/05000/the_positivism_paradigm_of_research.16.aspx

[52] University of Warwick, https://warwick.ac.uk/fac/soc/ces/research/current/socialtheory/maps/criticalrealism/#:~:text=Critical%20Realism%20(CR)%20is%20a,perceptions%2C%20theories%2C%20and%20constructions

E. **Realist closer to Holist** - Wider boundary conceptualised for the context-mechanism-outcome configurations; Uses mixed methods with greater role of the qualitative. Considers context and linkages with maximum of three of the following: Historical analysis, Epidemiological orientation, Knowledge pluralism and Bottom-up system vantage point;

F. **Holist** - 'Holism' is a term much used in relation to health. In relation to health knowledge, at the individual level, it means that the mind and the body are intrinsically intertwined, and the individuals are embedded in their ecological and social context. At the societal level, holism means that communities are not simply aggregates of individuals but are greater than the sum of the individuals, societies are not just aggregates of communities but greater than their sum due to their inter-relationships. Similarly, organizations are not an aggregate of their structure alone but of their value frames, the interactive processes among those who comprise them and their interactions with their social context.

**7. Types of Research**

A. **Evaluation research -** A research project that has as its focus the evaluation of some program process, policy or product. Unlike program evaluation, evaluation research is intended to generate knowledge that can inform both decision-making in other settings and future research.[53]

B. **Implementation Research -** Implementation Research (IR) is a form of enquiry into the process of translating clinical and public health evidence into practice. It addresses issues of improving access to and use of specific interventions within a local context of implementation.[54]

C. **Policy Analysis** – Policy Analysis is the process of identifying potential policy options that could address your problem and then comparing those options to choose

[53] Canadian Institute of Health Research, "A Guide to Evaluation in Health Research", https://cihr-irsc.gc.ca/e/documents/kt_lm_guide_evhr-en.pdf

[54] WHO, https://www.who.int/teams/health-ethics-governance/governance/health-systems-and-implementation-research#:~:text=Implementation%20Research%20(IR)%20is%20a,a%20local%20context%20of%20implementation

the most effective, efficient, and feasible one.[55]

D. **Research/Scientific Synthesis** - Research or scientific synthesis is the integration and assessment of knowledge and research findings pertinent to a particular issue with the aim of increasing the generality and applicability of, and access to, those findings.[56]

**8. Nature of Disciplinary Interaction**

i. **Multidisciplinarity** - Multidisciplinarity draws on knowledge from different disciplines but stays within the boundaries of those fields.[57]

ii. **Interdisciplinarity** - Interdisciplinarity analyzes, synthesizes and harmonizes links between disciplines into a coordinated and coherent whole.[58]

iii. **Transdisciplinarity** - Transdisciplinarity integrates disciplinary and lay insights towards co-production of knowledge and in so doing transcends traditional disciplinary boundaries.[59]

---

[55] Centers for Disease Control and Prevention, https://www.cdc.gov/policy/polaris/policyprocess/policyanalysis/index.html

[56] Hampton, S.E. & Parker, J.N. (2011) Collaboration and Productivity in Scientific Synthesis, BioScience, 61(11), 900–910, https://doi.org/10.1525/bio.2011.61.11.9; Magliocca, N. R., et.al (2015). Synthesis in land change science: methodological patterns, challenges, and guidelines. Regional environmental change, 15, 211-226; Baron, J.S., et.al (2017). Synthesis Centers as Critical Research Infrastructure, BioScience, 67(8), 750–759, https://doi.org/10.1093/biosci/bix053

[57] Choi, B.C.K & Pak, A.W.P. (2006). Multidisciplinarity, interdisciplinarity and transdisciplinarity in health research, services, education and policy: 1. Definitions, objectives, and evidence of effectiveness. Clinical and investigative medicine, 29(6), 351-64. https://pubmed.ncbi.nlm.nih.gov/17330451/

[58] Choi, B.C.K & Pak, A.W.P. (2006). Multidisciplinarity, interdisciplinarity and transdisciplinarity in health research, services, education and policy: 1. Definitions, objectives, and evidence of effectiveness. Clinical and investigative medicine, 29(6), 351-64. https://pubmed.ncbi.nlm.nih.gov/17330451/

[59] Choi, B.C.K & Pak, A.W.P. (2006). Multidisciplinarity, interdisciplinarity and transdisciplinarity in health research, services, education and policy: 1. Definitions, objectives, and evidence of effectiveness. Clinical and investigative medicine, 29(6), 351-64. https://pubmed.ncbi.nlm.nih.gov/17330451/
