## Supplementary 4: Application of the DHSRF to Review Studies: Some Illustrations for "The Dialogic Health Systems Research Framework (DHSRF): A tool for facilitating self-criticality, researcher interactions and knowledge management in Health Systems Research & Policy Studies"

### Study 1

| a) Description of the Study 1 | |
|---|---|
| **Name of study** | Does exposure to health information through mobile phones increase immunization knowledge, completeness and timeliness in rural India? |
| **Year of publication** | 2021 |
| **Author/s** | Arpita Chakraborty, Diwakar Mohan, Kerry Scott, Agrima Sahore, Neha Shah, Nayan Kumar, Osama Ummer, Jean Juste Harrisson Bashingwa, Sara Chamberlain, Priyanka Dutt, Anna Godfrey, Amnesty Elizabeth LeFevre, On behalf of the Kilkari Impact Evaluation Team |
| **Institutions (Author Affiliations)** | Statistics, Evidence, Accountability Programme, Oxford Policy Management, New Delhi, India; Department of International Health, Johns Hopkins Bloomberg School of Public Health, Baltimore, Maryland, USA; BBC Media Action India, New Delhi, India; Computational Biology Division, Department of Integrative Biomedical Sciences, Institute of Infectious Disease and Molecular Medicine (IDM), University of Cape Town Faculty of Health Sciences, Cape Town, South Africa; BBC Media Action, London, UK; University of Cape Town School of Public Health and Family Medicine, Cape Town, South Africa |
| **Funding** | Bill and Melinda Gates Foundation |
| **Type of Institution** | Global Development Consultancy, Multilateral Organisation, Academic/Research Institution |
| **Type of Research / Purpose** | Evaluative Research / To assess the determinants of parental knowledge of immunization and timely immunization in the context of the Kilkari programme. |
| **Objective of the study** | To explore determinants of parental immunization knowledge and immunization practice (completeness and timeliness) for children 0–12 months of age from four districts (Rewa, Hoshangabad, Mandsaur and Rajgarh) in Madhya Pradesh. |
| **Methodology of the study** | Descriptive & Analytical |
| **Level of analysis (Global/Inter-state/National/State/District)** | Districts (four) |
| **Area of analysis (Urban/rural/peri-urban/All)** | Rural |

| Policy Implications of the Study | The study provides quantitative insights into the effect of the largest direct-to-beneficiary mobile communication (Kilkari) on the immunization knowledge and practices of parents - completeness and timeliness of vaccinations in rural context of Madhya Pradesh (four districts). From the study results, we find that the programme had a limited impact on parental knowledge and practices. The study provides limited insights for policy makers in terms of recommendations directed at improving the content and approach of communication strategies in mHealth only. An examination of reasons behind: the increased knowledge levels in fathers that did not translate into timely and complete immunizations, the lack of improvement in mothers' knowledge, the possibility of inadequacies in coverage and quality of services or in the content of the messages, etc. through a qualitative inductive approach (situated in the Realist/Holist paradigm) would have provided more evidence and guidance for comprehensive policy-making and programme formulation. |
|---|---|

This evaluative quantitative descriptive research seeks to assess the determinants of parental knowledge of immunization, full immunization and timely immunization in the context of the Kilkari programme in rural contexts of four districts of Madhya Pradesh. This research is limited to data collection from mothers of children 0-12 months of age with access to mobile phones and their spouses. Using descriptive and inferential statistics, the study finds gaps in knowledge, timeliness and completeness for immunization for children in this rural context. The researchers suggest greater parental engagement with the help of intensified immunization content and a mechanism to enrol both spouses as well as a structured and focused approach to reach socially and economically vulnerable populations in remote areas.

This study is assigned to the Positivist paradigm as it tries to measure the association between socio-economic-demographic factors and the impact of mobile messaging intervention on parents' immunization knowledge and practices. The researchers have undertaken a quantitative operational approach and do not incorporate epidemiological or historical immunization analysis in these districts' rural context. While the researchers acknowledge the utility of mobile messaging programmes as one source of information on immunization, they present an analysis showing Kilkari's limitations - in terms of improving the father's knowledge but not the mothers. However, the study fails to go in-depth into the reasons behind the quantitative findings in the context of these study and looks at the available literature to interpret/qualify these findings.

The study's boundary is limited to addressing social determinants of health and the formal health service system, particularly on information systems and community engagement for immunization. While aspects of the socio-political contexts of health care of the people are gathered, the focus is largely on establishing quantifiable associations between these aspects and the immunization knowledge and practices of the parents. This research has a top-down health system vantage point as it studies an expert-formulated intervention (Kilkari), and its impact on the parents' knowledge and practices of immunization. The study is also conceived and undertaken from the researchers' perspective. The research acquires a positivist nature through its quantitative methodological approach without epidemiological orientation or historical analysis of immunization in the study areas and an absence of exploring the respondents' perceptions about the intervention or reasons they may have for listening/not listening or following/not following the advice given over phone as per the intervention. Nor do they examine the differences in health service coverage and quality across the districts by asking the parents or the service providers. While researchers are conscious of the rural context within which the study is situated in, they do not examine the socio-economic-demographic and cultural factors that govern mothers' knowledge and immunization practices despite the reach of the mobile messaging intervention. They do not investigate the public discourse on immunization within these contexts, informal health services, or socio-economic and cultural norms that deter timely and complete immunization. The study recommendations are limited to improving the content and reach of the intervention rather than finding solutions to resolve problems within the children's socio-political contexts of health care. Incoherence exists between the findings and the recommendations in terms of the gaps in the former that informed the latter. The authors do not examine the reasons for non-listening to messages or the reason why, even after listening to mHealth messages by fathers (and its resultant increase in the immunization knowledge in them), the completion and timeliness behaviour (of immunization) has not improved. It may be due to issues with the coverage and quality of services, or the inadequacy in the content of messages or real-life constraints of the parents, etc. Thus, in research such as this, which seeks to understand the impact of an intervention on people's knowledge and behaviour, an inductive qualitative exploration (in a Realist/Holist paradigm) would have additionally answered these questions and thereby provided a better programme and policy guidance.

Coherence between the research objectives, methods, results and recommendations of the study along with strengths of the research are detailed below.

| b) Assessment of Research Coherence | | | | |
|---|---|---|---|---|
| **Name of the Study** | **Health Systems Research Components** | | **Research Coherence** | **Strengths of research** |
| **Chakraborty, et.al (2014)** | **Research Objective(s)** | To explore determinants of parental immunization knowledge and immunization practice (completeness and timeliness) for children 0–12 months of age from four districts in Madhya Pradesh. | The study conceptualisation and methodology with a top-down approach are coherent with the study objectives. The mentioned values/principles underlie the research objective, methods and findings. | 1. This study is a good example of an exploratory research on a technological intervention (Kilkari) to see if it has influenced immunization outcomes in a rural context. The study also presents the strengths as well as limitations of the intervention in an unbiased manner<br><br>2. The researchers have focused on the measurable aspects of factors that influences immunization outcomes after the intervention as per the objectives of the research. |
| | **Research Question(s)** | N/A | | |
| | **Values & Principles** | Sustainability in social terms and in health outcomes, Equity, Context Appropriateness, Effectiveness, Appropriate Technology, Autonomy, Empowerment, Limited Trust and Responsiveness | | |
| | **Health System Problem Conceptualization** | The study is conceptualised as an exploratory exercise to fulfil the study objective. | | |
| | **Methodology** | The study design is quantitative in nature involving a cross-sectional survey data captured as part of a randomised controlled trial (RCT) of the Kilkari programme in four districts of Madhya Pradesh, India. Data was collected from 4423 postpartum women with a live birth during the index pregnancy and their husbands (3781 men - those who had not migrated for work). Information collected on the variables such as socio-economic status, mother's decision-making capacity in the household, and on her pregnancy, caste status of the ASHA worker is used for quantitative analysis, without deriving any qualitative information explaining the quantitative results. The study is largely top-down in nature as it is conceived and | | |

| | | | |
|---|---|---|---|
| | | undertaken by the researchers. | |
| | **Findings** | The study found that exposure to Kilkari immunization calls was associated with higher mean immunization knowledge among men but not women. The overall timeliness of immunizations received to date was 6% and 1% for basic and comprehensive packages, respectively. 47% of children 0–12 months old had been fully vaccinated<br>for the basic package at 12 months of age, around 44% had received the full comprehensive package.<br><br>These findings inform us that the programme did not translate into improved parental immunization practices as expected.<br><br>They have also estimated the association between different socio-economic variables and immunization uptake. While the actual qualitative reasoning behind these findings is not dealt with in detail in the study, the researchers refer to the literature that reports similar findings. For instance, the finding that exposure to Kilkari increased men's but not women's immunization knowledge is linked to men's higher access to mobile phones in India as noted in Barboni, et.al (2018). | The recommendations are broadly based on the quantitative results of the study, and literature review by the researchers. The study does not examine the reasons behind the differing levels of reach of the messages between fathers and mothers, non-listening to messages, or why after listening to these messages and increased knowledge among fathers, there is a failure in improving completion and timeliness behaviour of immunization. The results also do not seem to hint at large improvements in immunization outcomes because of the intervention.<br><br>The study recommendations focus only on the content and approach in communication to parents and those residing in remote locations, ignoring the larger socio-political-economic-cultural factors that ultimately determines parental decision-making, completion and timeliness of their child's immunization. As stated previously, though some of these factors were included in the quantitative analysis, the study fails to go for an in-depth qualitative exploration of these factors in this context. |

| | | | |
|---|---|---|---|
| | **Recommendations** | 1. Intensified immunization content, and a mechanism to enrol both spouses whenever possible, may achieve deeper impact. Further efforts to use mobile-based communication to raise awareness about the benefits of immunization, supported by mass media communication, is even more crucial with the backdrop of COVID-19.<br>2. Structured and focused approach needs to be adopted to reach the socially and economically vulnerable in remote areas, specifically for immunization practice. | Therefore, incoherence exists between the findings and recommendations in this study |

### *Application of Tools*

The following describes the values and principles underlying the conceptualization of health systems, elements of conceptualization of research, and methodological approach of this publication using the newly developed Health Systems Research Framework. These are the steps for assessing the research coherence of this study.

#### Step 1. Analysis of Values and Principles informing Health Systems conceptualisation

| **Name of the Study** | | | **Chakraborty, et.al (2014)** | |
|---|---|---|---|---|
| | **Values & Principles** | **Values & Principles relevant for the purpose of the study (Yes/No)** | **Absent/Limited/ Present** | **Observations** |

| | | | | |
|---|---|---|---|---|
| **Health System goals** | **Sustainability** | Yes | Present | Sustainability in Social and Health Outcomes terms:<br>The study seeks to understand the impact of a maternal messaging programme on the immunization knowledge and practices in rural MP, and how it needs to be improved accounting for the socio-economic disparities. |
| | **Equity** | Yes | Present | The study notes the influence of caste, educational attainment of parents, employment status of mothers, wealth on parental knowledge and uptake of immunization. Families with more financial resources and whose mothers had more time in the home may find it easier to access health facilities on time for immunization as compared with poorer families, families where the mother worked and families with many children. |
| | **Context appropriateness** | Yes | Present | The study was conducted in rural Madhya Pradesh which has one of the lowest per capita state domestic products in India and has reported significant gender gap in women's literacy and access to mobile phones within the state. The state falls behind (below the national averages) in health indicators including immunizations. |
| | **People-centredness** | Yes | Absent | N/A |
| **Health System functioning** | **Effectiveness** | Yes | Present | Exposure to Kilkari calls was associated with higher likelihood of children 0–12 months receiving birth vaccines on time but had no association with overall full and timely immunization for the basic or comprehensive immunization package. The study reveals the percentages of children vaccinated for the basic and comprehensive packages, and the overall timeliness in receiving these packages. While the Kilkari backend data show whether a Kilkari call was answered and whether it was allowed to play, it cannot reveal who picked up the call and who listened to the message. |
| | **Safety** | No | Absent | N/A |
| | **Clinical Rationality** | No | Absent | N/A |
| | **Ethical Practice** | Yes | Absent | N/A |
| | **Appropriate Technology** | Yes | Present | The study discusses the impact of Kilkari programme on parental knowledge and practices of immunization for their children in a rural context. |

| | | | | |
|---|---|---|---|---|
| | **Technical efficiency** | Yes | Absent | N/A |
| | **Affordability** | Yes | Absent | N/A |
| | **Ecological sensitivity** | No | Absent | N/A |
| | **Dignity in care** | Yes | Absent | N/A |
| | **Self-reliance** | No | Absent | N/A |
| | **Autonomy** | Yes | Present | The study seeks information on the autonomy of parents who decide whether to get their children vaccinated or not after receiving immunization related messages via Kilkari programme. It also assesses the percentage distribution of decision-making power in households with respect to daily purchases, pregnancy and number of children among men and women. |
| | **Empowerment** | Yes | Present | The study notes how parents are empowered with the necessary information for immunization uptake of their children. |
| | **Trust** | Yes | Limited | Trust in the health service system is not explored in depth in this study. While the study discusses how people grasp information provided via the Kilkari programme and how far that has translated into their immunization knowledge and practices, the researchers do not look into the reasons behind delayed or incomplete immunization, and whether this has something to do with trust of the parents on their local health service systems. |
| | **Transparency** | No | Absent | N/A |
| | **Accountability** | No | Absent | N/A |
| | **Responsiveness** | Yes | Limited | The study discusses how the Kilkari programme was undertaken in the rural context of Madhya Pradesh and observes that the exposure was not associated with overall improvements of full and timely immunization coverage. Hence, the researchers recommend further efforts to ensure that socially and economically vulnerable people in remote areas, both spouses in a household, are adequately reached to achieve deeper impact. |
| | **Decentralisation (dialogic, deliberative, democratic)** | Yes | Absent | N/A |

### Step 2. Analysis of Conceptualisation of the Health System

<table>
<tr><td colspan="3">Name of the Study</td><td colspan="3">Chakraborty, et.al (2014)</td></tr>
<tr><td colspan="3">Conceptual framework</td><td colspan="3">Not Stated</td></tr>
<tr><td colspan="3"></td><td>Components relevant for the purpose of the study</td><td>Absent/Limited/ Present</td><td>Observations</td></tr>
<tr><td rowspan="5">Health System Conceptualisation</td><td colspan="2">Boundary</td><td>Addressing Social and Ecological Determinants of Health,<br>Formal Health Service System</td><td>Limited</td><td>Addressing Social Determinants of Health, Formal Health Service System</td></tr>
<tr><td colspan="2">Subsystems of formal health service system (Structures and processes)</td><td>Health Service delivery, Infrastructure, Health workforce, Health Financing, Health Governance, Information Systems, Access to essential medicines, vaccines, Community Engagement - Organization and operationalization</td><td>Limited</td><td>Information Systems, Community Engagement - Operationalization</td></tr>
<tr><td colspan="2">Informal Health Services (Structures and processes)</td><td>N/A</td><td>Absent</td><td>N/A</td></tr>
<tr><td colspan="2">Health System Vantage point</td><td>Combined</td><td>Limited</td><td>Top-Down</td></tr>
<tr><td>Dynamic Elements of Health System and its context</td><td>Ecosystems</td><td>Yes</td><td>Absent</td><td>N/A</td></tr>
</table>

| | | | | | |
|---|---|---|---|---|---|
| | | **Socio-political contexts of health care** | Yes | Limited | The researchers have gathered information on parental socio-economic-demographic, educational and employment characteristics, women's role in decision-making in daily purchases, pregnancy, number of children, and their access to mobile phones, sources of immunization information, etc. in this rural context. However, this information was used in quantitative analysis to measure the influence of these factors on parental immunization knowledge and uptake. There is an absence of a detailed investigation into how these factors have influenced access to immunization services and their uptake in a qualitative manner in the study. |
| | | **Meaning systems of health and health care** | Yes | Absent | N/A |
| | | **Informal social arrangements & Community practices for health** | Yes | Absent | N/A |
| | **Relationship of the elements across the system** | | | Absent | N/A |
| | **Theory of change** | | Assessing the impact of a mobile messaging service to provide information on child immunization to parents to see how far their knowledge and practices have improved will allow identification of challenges and the need for structured and focused correctives to reach everyone, especially the socially and economically vulnerable people in remote contexts, and women whose decision-making power within households are constrained by societal/cultural norms. | | |

### Step 3. Analysis of Methodological approaches

| **Name of the Study** | **Chakraborty, et.al (2014)** |
|---|---|

| Methodological Approaches to Health Systems Research | | Methodological Approaches relevant for the purpose of the study | Absent/Limited/ Present | Observations |
|---|---|---|---|---|
| **Analytical approaches** | **Epidemiological approach (Health profile and determinants)** | Yes | Absent | N/A |
| | **Historical analysis** | Yes | Absent | N/A |
| | **Social Science approaches** | Yes | Limited | While the study notes the influence of social determinants of health on immunization uptake in quantitative terms (odds ratio and statistical tests of significance), it does not qualitatively investigate into how these factors - such as women's low decision-making power within households on pregnancy and number of children, access to mobile phones, employment status, etc. - translates into immunization outcomes for their children. |
| | **Management Science approaches** | No | Absent | N/A |
| | **Ecosystem approaches** | No | Absent | N/A |
| **Operational approaches** | | Non-Intervention Study Design – Mixed Methods | Non-Intervention Study Design - Quantitative | A quantitative study design was used to explore determinants of parental immunization knowledge and immunization practice (completeness and timeliness) in four districts in Madhya Pradesh. This study is part of a larger RCT to assess the impact of Kilkari on immunization outcomes in rural MP. The main data sources included a cross-sectional survey and vaccination cards. Quantitative analysis of the data was undertaken using ordered logistic regressions to analyse the factors associated with parental immunization knowledge, and a Heckman two-stage probit model to analyse completeness and timeliness of immunization after correcting for selection bias from being able to produce the immunization card. |

| | | | |
|---|---|---|---|
| **Nature of disciplinary interaction** | Interdisciplinary | Interdisciplinary | The study utilises knowledge from biostatistics, public health and limited socio-economic analysis to analyse the factors associated with parental immunization knowledge, completeness and timeliness of immunization after the introduction of Kilkari programme. Knowledge from these disciplines is used in a coordinated and coherent manner, without creating silos of information drawn from these disciplines. |
| **Ethical Considerations of the Study** | 1. Informed Consent: Verbal informed consent was obtained from the study participants, for participation in the RCT and in the baseline and postpartum survey.<br>2. Confidentiality: None mentioned<br>3. Conflicts of interest: The authors declare that they have no competing interests. | | |
| **Quality Assessment of Research Operationalisation*** | Methodological Quality: High<br>The researchers have detailed the quantitative approach, data collection methods with justifications in this study. Findings have been adequately derived from the data via ordered logistic regression and a Heckman two-stage probit model. Results are interpreted from these quantitative analyses and literature reviews. Coherence exists between data sources, data collection, and results in the study. | | |
| **Research paradigm** | Positivist | | |

*** Using the Mixed Methods Appraisal Tool adapted from Hong, et.al (2018)**

Thus, this study illustrates the limitations of well-conducted research in the Positivist paradigm when the objective involves exploring the impact of an intervention on parental knowledge and practices of immunization in rural contexts. From this study's results, it is understood that merely imparting knowledge about immunization is not an adequate strategy to improve timely uptake of the services. Next, we shall go through an illustration of a study which adopted a Holist paradigm.

**Study 2**

| **a) Description of the Study** | |
|---|---|
| **Name of study** | Advancing the application of systems thinking in health: understanding the growing complexity governing immunization services in Kerala, India |
| **Year of publication** | 2014 |
| **Author/s** | Joe Varghese, V Raman Kutty, Ligia Paina & Taghreed Adam |

| | |
|---|---|
| **Institutions (Author Affiliations)** | Centre for Chronic Disease Control and Governance Hub, Public Health Foundation of India; Achutha Menon Centre for Health Science Studies, Sree Chitra Tirunal Institute for Medical Science and Technology, Thiruvananthapuram, India; Department of International Health, Johns Hopkins University School of Public Health, Baltimore, USA; Alliance for Health Policy and Systems Research, World Health Organization, Geneva, Switzerland |
| **Funding** | International Development Research Centre, Ottawa, Canada. |
| **Type of Institution** | Academic/Research Institution |
| **Type of Research / Purpose** | Exploratory Research / To understand the complexity in governance of immunization services using complex adaptive systems lens |
| **Objective of the study** | To describe an application of complex adaptive systems theory and methods to understand and explain the phenomena underlying unexpected changes in vaccination coverage |
| **Methodology of the study** | Explanatory |
| **Level of analysis (Global/Inter-state/National/State/District)** | Districts |
| **Area of analysis (Urban/rural/peri-urban/All)** | Not specified |
| **Policy Implications of the Study** | This study discusses the utility of a complex adaptive systems lens to systematically explore the driving forces and factors in each setting and develop appropriate and timely strategies to address them. The study calls for greater consideration of dynamics of vaccine acceptability while formulating immunization policies and program strategies. Public health governance should account for not just epidemiological and economic analyses, but also, the nature of multiple interactions between the public health department, providers, social networks and households on a public health function such as immunization. Their perceptions and ideas shaped by trust and the complexity of these interactions that determine response to immunization are too crucial to be missed. Therefore, the study espouses the importance of values of trust, autonomy (of beneficiaries), acceptability (of public health initiative), and responsiveness (of officials to the needs and perceptions of the community) in policy making for improving people's trust in the health service system and thereby enhanced uptake of services such as immunization. |

This exploratory case study research describes an application of complex adaptive systems (CAS) theory and methods to explain the phenomena underlying unanticipated variations in immunization coverage. This research is not limited to allopathic practitioners or community health workers, but also accounts for the views of those who oppose vaccinations. The study finds changes in average immunization across districts due to variations in the levels of trust – interpersonal and institutional. The role of trust in health workers and institutions shaping interactions among different actors is expressed via causal loop diagrams. Apart from suggesting the utilization of this theory and its methods in immunization research, the researchers recommend greater consideration of dynamics of vaccine acceptability while formulating immunization policies and program strategies including pluralistic dialogue with other systems of medicine drawing from the qualitative results.

This qualitative research study is assigned to the Holist paradigm as it adopts the CAS lens for a complex problem of uncovering the real drivers behind the unanticipated variations in immunization coverage in two districts of Kerala, drawing from various sources of knowledge, and thereby addressing the coverage declines. The study has a wide configuration of socio-political contexts, public discourse surrounding the immunization programme (in detail), experience of immunization services on prevention of childhood diseases, individual and community perceptions of immunization and the programme. The research acquires a holistic nature via its methodological approaches involving:

- **Historical analysis** – The study presents the Acceptability and Vaccine-Resistance Phases to Immunization Programme in Kerala, how authorities responded to the problem of decline in immunization coverage and discusses the impact of their response in the presence of certain actors who have a disproportionate influence over household's vaccination decision.

- **Knowledge pluralistic approach** – The study accounts for the views of those who propound and those who oppose immunizations. These conversations describe the underlying politics of knowledge (between systems of medicine) that influences their response to pulse polio initiative, and effects on community's perceptions. For instance, the alternative system of medicine practitioners mention that their response against IPPI arose from: one, the superiority shown by allopathic practitioners and lack of recognition given to the alternative systems, and two, the public health administration (largely consisting of allopathic practitioners) retaliated against homoeopathic professionals (when the immunization coverage started decreasing) by issuing a government order to set-up vaccine booths in government owned homoeopathy dispensaries. These nuances of power and hierarchy within systems of medicine are brought out which could reason why the immunization coverages declined in North Kerala.

- **Epidemiological orientation –** The researchers have selected a high coverage and a low coverage district for the study, historically traced coverage, compared between the low coverage with high coverage districts, and point out that how deaths from a school vaccination program in the low coverage district further affected vaccine uptake in the northern part of Kerala.

- **Mix of top-down and bottom-up approaches –** The health system vantage point of this study combines both institutional and community approaches wherein the perspective underlying immunization policymaking is found to be not just dependent on planning by policy-makers and providers, but also on household perception of other actors' interests, exercising of autonomy of parents/beneficiaries and the role of informal interactions that determines trust in vaccines.

Further, the researchers have attributed great importance to the value of contextuality wherein they have utilized an observation guide to gather insights into cultural meanings and interpretations related to provider and beneficiary. They examined the role of public discourse and its influence. The study findings and recommendations draw upon contextual inter-linkages between the actors and suggest engaging in dialogue to build consensus within the boundary of immunization services and public discourse. Through its methodology, the study has highlighted the utility of systems dynamics and Causal Loop Diagrams, and their flexible usage in planning and undertaking research based on local context.

Coherence between the research objectives, methods, results and recommendations of the study along with strengths of the research are detailed below.

| **b) Assessment of Research Coherence** | | | |
|---|---|---|---|
| **Name of the Study** | **Health Systems Research Components** | **Research Coherence** | **Strengths of research** |

| | | | | |
|---|---|---|---|---|
| **Varghese, et.al (2014)** | **Research Objective(s)** | To describe an application of Complex Adaptive Systems (CAS) theory and methods to understand and explain the phenomena underlying unexpected changes in vaccination coverage in two districts in Kerala, one with low coverage and the other with high coverage. | The study conceptualization and methodology with a combination of top-down and bottom-up approaches were coherent with the objectives of the study (to understand the reasons behind unanticipated changes in immunization coverage in the two districts). The mentioned values/principles underlie the research objective, methods and findings. | 1. The researchers have adopted the CAS lens for understanding and decoding a complex problem.<br><br>2. By using a bottom-up approach (perceptions of the community of the research problem) with politics of knowledge and knowledge pluralism (that informs community and other stakeholder responses/perceptions), and historical trajectory, the researchers are able to capture deeper insights into the factors that underlie the problem. |
| | **Research Question(s)** | N/A | | |
| | **Values & Principles** | Sustainability in terms of Acceptability of the Immunization Programme, Context Appropriateness, Effectiveness, Autonomy, Trust, Accountability, Responsiveness | | |
| | **Health System Problem Conceptualization** | Adopts the CAS lens for a complex problem of uncovering the real drivers behind the unanticipated variations in immunization coverage in two districts of Kerala, drawing from various sources of knowledge, and thereby addressing the coverage declines. | | |

| | | | |
|---|---|---|---|
| | **Methodology** | The study design is qualitative in nature with the Acceptability Phase and Vaccine-Resistance Phase to Immunization Programme in Kerala presented using causal loop diagrams indicating how the authorities responded to this problem and the impact of their response. The study accounts for the views of those who advocate and those who oppose immunizations and has a limited epidemiological orientation. With a mix of top-down and bottom-up approaches, the study demonstrates how the perspective underlying immunization policy making is not just dependent on planning by policymakers and providers, but also on household perception of other actors' interests, exercising of autonomy of parents/beneficiaries and the role of informal interactions that determines trust in vaccines. | |
| | **Findings** | The study found variations in coverage owing to issues with interpersonal and institutional trust mediated by externally driven and experientially informed public discourse. | The role of 'trust' and its determinants is arrived at through this qualitative research, bringing forth the recommendations such as engage in dialogue with groups that oppose immunization and utilize their expertise to put out consistent health messages, utilize the potential of right kind of interactions of ASHAs and Anganwadi Workers with households as well as experiences of positive benefits of immunization to reinforce trust, etc.<br>Thus, the study exhibits |

| | | | |
|---|---|---|---|
| | **Recommendations** | 1. Consistent health messaging from different sources of expertise and engaging in dialogue that generates consensus with other systems of medicine.<br>2. Utilize community level functionaries like ASHAs to influence household level decisions on immunization.<br>3. The evidence base of public health programs should go beyond epidemiological and economic analysis and therefore, there is a need for public health governance systems to take the nature of multiple interactions into consideration when societies organize themselves to manage provision of a public service like immunization. | coherence from its objectives to recommendations, moving across individual and collective perceptions to societal values and interactions within their context that influence their immunization decisions. |

#### *Application of Tools*

The following describes the values and principles underlying the conceptualization of health systems, elements of conceptualization of research, and methodological approach of this publication using the newly developed Health Systems Research Framework. These are the steps for assessing the research coherence of this study.

#### Step 1. Analysis of Values and Principles informing Health Systems conceptualisation

| **Name of the Study** | | | **Varghese, et.al (2014)** | |
|---|---|---|---|---|
| | **Values & Principles** | **Values & Principles relevant for the purpose of the study (Yes/No)** | **Absent/Limited/ Present** | **Observations** |
| **Health System goals** | **Sustainability** | Yes | Present | Acceptability of the programme: The study seeks to understand how peoples' acceptance/resistance of the immunization programme influences its |

| | | | | |
|---|---|---|---|---|
| | | | | uptake in the selected districts across the years. |
| | **Equity** | Yes | Absent | N/A |
| | **Context appropriateness** | Yes | Present | The study involved identification of different locations within the same district (low and high immunization coverage areas) helped in collecting information from diverse contexts. Participant and non-participant observations were made with the help of an observation guide to gather insights into cultural meanings and interpretations related to provider and beneficiary behaviours and context. |
| | **People-centredness** | Yes | Absent | N/A |
| **Health System functioning** | **Effectiveness** | Yes | Present | The analysis of trends in immunization coverage in both districts showed a sudden decline in immunization coverage in Kozhikode; based on three rounds of the District Level Household and Facility Survey, Kozhikode showed a decline after the second round of the survey in the 2002–2004 period. The full immunization coverage in Kozhikode district in northern Kerala dropped from 94% (2002–2004) to 65% (2007–2008). During the same period, the coverage in a southern district, Alappuzha, had in fact gone up from about 84% to around 92%. |
| | **Safety** | Yes | Absent | N/A |
| | **Clinical Rationality** | Yes | Absent | N/A |
| | **Ethical Practice** | Yes | Absent | N/A |
| | **Appropriate Technology** | No | Absent | N/A |
| | **Technical efficiency** | Yes | Absent | N/A |
| | **Affordability** | Yes | Absent | N/A |
| | **Ecological sensitivity** | No | Absent | N/A |
| | **Dignity in care** | Yes | Absent | N/A |
| | **Self-reliance** | No | Absent | N/A |
| | **Autonomy** | Yes | Present | The study observes the autonomy of beneficiaries who decide whether to be vaccinated or not. |
| | **Empowerment** | Yes | Absent | N/A |

| | | | | |
|---|---|---|---|---|
| | **Trust** | Yes | Present | The study notes the following with respect to trust on the immunization programme:<br>In the first view, acceptability to immunization in the initial phase in Kerala can be viewed because of the trust in institutions of professional expertise (in this case, medical knowledge). However, the conflicting messages that emerge from different systems of medicine challenge the trust which people attribute to expert systems of vaccination.<br>In the second interpretation, trust is approached as a cognitive phenomenon, or a judgement based on a rational gamble and therefore household perception of other actors' interests are important.<br>Further, trust in health workers can also be explained through the notions of 'affective trust', which is developed through emotional bonds and obligation generated through their repeated personal interactions with the households. |
| | **Transparency** | Yes | Absent | N/A |
| | **Accountability** | Yes | Present | The study calls for sensitising the media for more responsible reporting and using it to convey appropriate health messages are options that public health departments may use in such situations, even though it is unlikely to eliminate all unwanted information from reaching households. |
| | **Responsiveness** | Yes | Present | The study discusses how the authorities have responded to this problem of decline in immunization coverage and discusses the impact of their response in the presence of certain highly connected actors playing a disproportionate influence over a household's vaccination decision. |
| | **Decentralisation (dialogic, deliberative, democratic)** | Yes | Absent | N/A |

### Step 2. Analysis of Conceptualisation of the Health System

| | |
|---|---|
| **Name of the Study** | **Varghese, et.al (2014)** |
| **Conceptual framework** | **Complex Adaptive Systems Theory** |

<table>
<tr><th colspan="3"></th><th>Components relevant for the purpose of the study</th><th>Absent/Limited/ Present</th><th>Observations</th></tr>
<tr><td rowspan="6">Health System Conceptualisation</td><td colspan="2">Boundary</td><td>Addressing Social and Ecological Determinants of Health,<br>Formal Health Service System,<br>Informal Health Services</td><td>Present</td><td>Formal Health Service System</td></tr>
<tr><td colspan="2">Subsystems of formal health service system (Structures and processes)</td><td>Health Service delivery, Infrastructure, Health workforce, Health Financing, Health Governance, Information Systems, Access to essential medicines, vaccines, etc. - Organization and operationalization</td><td>Present</td><td>Health service delivery, Health governance, Public Discourse</td></tr>
<tr><td colspan="2">Informal Health Services (Structures and processes)</td><td>N/A</td><td>Absent</td><td>N/A</td></tr>
<tr><td colspan="2">Health System Vantage point</td><td>Combined</td><td>Present</td><td>Combined</td></tr>
<tr><td rowspan="2">Dynamic Elements of Health System and its context</td><td>Ecosystems</td><td>Yes</td><td>Absent</td><td>N/A</td></tr>
<tr><td>Socio-political contexts of health care</td><td>Yes</td><td>Present</td><td>The researchers have tried to understand the role of social, religious, and political (provider associations) factors that influenced the community and generated opposition against polio immunization in one of the selected districts of the study. They used a observation guide to understand the cultural meanings and interpretations<br>related to provider and beneficiary behaviours and context in this study.</td></tr>
</table>

| | | | | | |
|---|---|---|---|---|---|
| | | **Meaning systems of health and health care** | Yes | Present | The study considered views of immunization service providers from public and private sector, those who facilitate vaccination, such as community health workers, and those who opposed it, mothers of children below five years of age, community health workers, nutrition and pre-school teachers and community leaders. |
| | | **Informal social arrangements & Community practices for health** | Yes | Limited | The study is conceptualised to look at the utilization of immunization service in the selected districts. |
| | **Relationship of the elements across the system** | | | Present | N/A |
| | **Theory of change** | | Personal experience, a wider social experience and the public discourse around it shape the acceptance or resistance towards an intervention such as child immunization. The complexity of these interacting processes leads to emergent behaviour change that can either enhance or decrease vaccine uptake. The study finds that the measures required to build trust institutional and interpersonal trust are crucial to increase vaccination uptake. The social relationships and their dynamics in the community will affect how communitisation programmes will get implemented. | | |

### Step 3. Analysis of Methodological approaches

| **Name of the Study** | | **Varghese, et.al (2014)** | | |
|---|---|---|---|---|
| **Methodological Approaches to Health Systems Research** | | **Methodological Approaches relevant for the purpose of the study** | **Absent/Limited/ Present** | **Observations** |
| **Analytical approaches** | **Epidemiological approach (Health profile and determinants)** | Yes | Limited | The study considers a better performing and poor performing district in immunization coverages. |

| | | | | |
|---|---|---|---|---|
| | **Historical analysis** | Yes | Present | The researchers present the Acceptability Phase and Vaccine-Resistance Phase to Immunization Programme in Kerala. They discuss how the authorities responded to this problem of decline in immunization coverage and discuss the impact of their response in the presence of certain highly connected actors playing a disproportionate influence over a household's vaccination decision. |
| | **Social Science approaches** | Yes | Present | Knowledge Pluralism and Politics of Knowledge: The researchers interview those who promote and those who oppose immunizations. They bring forth the debates and conflicting views between practitioners of allopathy and alternate systems of medicine on the safety of immunizations that influenced the trust of the community in Northern Kerala. The study also shows how the allopathic system confronted the groups that opposed immunizations (homoeopaths) by getting a government order issued to set-up vaccine booths in government owned homoeopathic dispensaries. |
| | **Management Science approaches** | No | Absent | N/A |
| | **Ecosystem approaches** | No | Absent | N/A |
| **Operational approaches** | | Non-Intervention Study Design – Mixed Methods | Non-Intervention Study Design - Qualitative | A qualitative case study design was used to obtain an understanding of immunization coverage in Kerala. The main data sources included a literature and document review (including news reports), in-depth interviews, focus group discussions, and observations of immunization services. Qualitative analysis of the data was done via deductive coding. Triangulation of data was ensured across the three methods of data collection. Based on the qualitative data analysis, a CLD was developed to assist in the identification and interpretation of the feedback loops that emerged in the context of immunization. |

| | | | |
|---|---|---|---|
| **Nature of disciplinary interaction** | Interdisciplinary | Interdisciplinary | The study utilises knowledge from epidemiology, historical analysis, and politics of knowledge to analyse the reasons behind unanticipated changes in immunization coverages in the study areas in a coordinated and coherent manner, without creating silos of information drawn from these disciplines. |
| **Ethical Considerations of the Study** | 1. Informed Consent: Written permission for data collection was obtained from state level health officials as well as from district level officials and participation in the study was made voluntary by ensuring informed consent from all participants and the possibility to withdraw at any time.<br>2. Confidentiality: All identifiers of the study participants from the transcripts of the data were removed by the first author to ensure anonymity of the study participants.<br>3. Conflicts of interest: The authors declare that they have no competing interests. | | |
| **Quality Assessment of Research Operationalisation*** | Methodological Quality: High<br>The researchers have detailed the qualitative approach, data collection methods with justifications in this study. Findings have been adequately derived from the data via deductive coding. Results are interpreted using the Causal Loop Diagrams and supported with quotes from stakeholder interviews. Coherence exists between data sources, collection, analysis and interpretation in this study. | | |
| **Research paradigm** | Holist | | |

*** Using the Mixed Methods Appraisal Tool adapted from Hong, et.al (2018)**

### References to Supplementary 4

Chakraborty, A., Mohan, D., Scott, K., et al. (2021). Does exposure to health information through mobile phones increase immunization knowledge, completeness and timeliness in rural India? *BMJ Global Health*, 6 (e005489). doi:10.1136/bmjgh-2021-005489

Barboni, G., Field, E. & Pande, R. (2018). A Tough Call: Understanding the Barriers to, and Impacts of, Women's Cell Phone Adoption in India. MA, USA: Harvard Kennedy School, Evidence for Policy Design. https://epod.cid.harvard.edu/sites/default/files/2018-10/A%20Tough%20Call_2.pdf

Hong, Q. N., Pluye, P., Fàbregues, S., Bartlett, G., Boardman, F., Cargo, M., ... & Vedel, I. (2018). Mixed methods appraisal tool (MMAT), version 2018. *Registration of copyright*, *1148552*(10). Retrieved from: http://mixedmethodsappraisaltoolpublic.pbworks.com/w/file/fetch/127916259/MMAT_2018_criteria-manual_2018-08-01_ENG.pdf

Varghese, J., Kutty, V.R., Paina, L., Adam, T. (2014) Advancing the application of systems thinking in health: understanding the growing complexity governing immunization services in Kerala, India. *Health Research Policy and Systems, 12*(47). doi: 10.1186/1478-4505-12-47.
