## Supplementary 5: Illustration of Designing of HSR using the DHSRF for "The Dialogic Health Systems Research Framework (DHSRF): A tool for facilitating self-criticality, researcher interactions and knowledge management in Health Systems Research & Policy Studies"

Here, we detail the conceptual components related to the value/principle of technical efficiency to show how the DHSRF can help a researcher to think about different research questions and to what extent different paradigms shape the scope of such an exercise. This table is indicative and may include many more research questions and approaches. What is best suited for the research will have to be decided in coherence to the research objectives. In this hypothetical exercise of designing a HSR study for researching on the theme of immunization, we look at the three distinct research paradigms and see how they differ and even complement in conceptualising and operationalising the research questions.

**Application of the HSRF to design a study: Detailing of Technical Efficiency as a value**

| **Researcher's framework** | | | **Possible applications of research paradigms** | | |
|---|---|---|---|---|---|
| **Boundary** | **Conceptual elements and their possible research questions** | **Methodological approach(es)** | **Positivist** | **Realist** | **Holist** |
| SDH + Formal Service system and their sub-systems | Can consider the inclusion of a combined vantage point on planning and implementing immunization programmes, effects of immunization supply chain and infrastructure on service delivery, and thereby on overall population health. | Interdisciplinary approach | | | |
| | **Ecosystem** | | | | |

| | | | | | |
|---|---|---|---|---|---|
| | How does the local geography (terrain, distance and time) and environmental conditions determine access to timely immunization services?<br><br>How can the efficiency of the immunization supply chain be improved in difficult terrains and remote areas? | Ecosystems approach; Eco-social approach; Social mapping; GIS; Questionnaire based Survey, Historical analysis, Analysis of social stratification; Cost and time analysis etc. | The Positivist approach would more likely use GIS mapping techniques to estimate the costs, distance and time taken to reach nearest health facilities in different terrains. | A Realist approach will include questionnaires/ interviews with providers, supply chain personnel and community stakeholders in diverse geographical and environmental circumstances on their experiences and perceptions of the immunization delivery system. | The Holist would include all of the other two and, in addition, take a historical perspective of changes in geology, environmental conditions, infrastructural, transport and technological development over the years, peoples' changing perceptions towards vaccinations, and bottom-up difficulties and solutions of the local community (particularly of the marginalised sections) for accessing vaccination services. |
| | **Socio-economic context of health and health care** | | | | |
| | How do the differences in socio-economic contexts of communities' influence immunization service delivery and uptake? | Historical analysis; economic analysis; discourse analysis; political science approach; Sociological and anthropological approaches, cultural studies, etc. | Situation analysis of immunization programme implementation and population level socio-economic, demographic, education, and health indicators at a point of time, and assessment of changes over time. | Analysis of policy and planning documents about immunization programmes in India, analysis of the earlier mentioned *(previous column)* factors shaping the historical and political context in which the vaccination programme was conceptualised and continues to function, and the political economy of the immunization programme. | Like the realist approach with special focus on people-centeredness of services, equity, and affordability as well as people's perceptions and practices regarding prevention of children's ill-health and promotion of their health, and the place of immunization within that. The diversity of perceptions and behaviours across socio-economic sections would be understood historically to explain the present. |

| | | | | | |
|---|---|---|---|---|---|
| | How does the health system ensure efficiency in immunization service delivery in a particular geographical unit? | Economic analysis, political economy of health approach and management science | 1.Review of immunization programme documents.<br>2.Facility based survey to assess the status of immunization service delivery.<br>Survey and quantification to assess access of population across different economic and social groups to immunization, the coverage, and time taken in accessing services. | Analysis of public and private immunization services utilisation changes across the years and Particularly in different contexts (including analyses of the contexts); Community/user perceptions of immunization service uptake across health facilities in diverse contexts.<br>Assess where efficient service delivery is ensured and where it is not, and reasons behind the same. | Historical analysis of immunization service delivery in the specific geographical unit, Assess the epidemiological needs of the contexts, and private sector provisioning of immunization through textual analysis, health researchers and policy makers' interviews, in addition to the previous. Exploration of people's immunization seeking behaviour and practices in scenarios of differential access. |
| | How can efficiency be improved in vaccine development and introduction during a public health emergency? | Economic modelling; Epidemiology; Management science | Costing analysis of vaccine development roll-out; Input-output analysis; Cost-benefit analysis; Comparison of costs of vaccine introduction vis-à-vis reduction in disease burden. | Similar to the positivist, and including contextual views of policy makers, health administrators, vaccine developers/researchers and finance department, drug regulatory, public health officials on the subject. | In addition to previous, historical analysis of vaccine development, costs and funding. Assessment of costs to examine if that will align with the epidemiological, socio-economic, demographic needs, provider and community felt needs. Comparison of costing of vaccination based on current and future epidemiological and demographic trends and the current status of public funding/income levels of the community. |
| | **Meaning systems** | | | | |

| | | | | | |
|---|---|---|---|---|---|
| | How do providers perceive, plan and undertake efficient immunization services in a specific geographical unit? | Sociology, Political science; Management science; Politics of Knowledge | Quantification of perceptions of providers of different systems of medicine; Input-output analysis, time and costs incurred in immunization service delivery against optional interventions such as for environmental control or raising immunity | Qualitative exploration of perceptions of different providers across different positions and systems of medicine, assessment of their perceptions vis-a-vis power relations in different contexts. The optional preventive measures are likely to be more diverse. | In addition to the realist, historical analysis of immunization services, perceptions of different systems of medicine towards immunization, and their impact on community uptake of the service (from institutional perspective). |
| | How do people perceive and utilize immunization services in a specific geographical unit? | Same as above | Quantified assessment of people's perceptions, time and costs incurred in uptake of immunization services. | Mixed methods exploration of people's perception across different socio-economic sub-groups as well as the social context of the specific locations. Thereby understanding and explaining the diverse perceptions and behaviours. | In addition to the previous two, historical assessment of immunization services, impact of complementary/alternativ e systems of medicine on immunization uptake by the community (from bottom-up perspective), in different contexts; community perceptions of what all is required for improving their health and the place of immunization within it. |
| | **Informal Social arrangements & Community practices for health** | | | | |
| | Has immunization become a people's practice and health seeking behaviour that is actively sought? | Sociology; Anthropology; Epidemiology; Politics of Knowledge; Management science | Trends in utilisation of immunization services across geographies. | Trends in utilisation of immunization services across geographies, socio-economic groups and communities; also exploring the reasons for the differentials. | A historical and bottom-up assessment of community perceptions of all that is required for improving their health and the place of immunization within it. Also, what is the active process of seeking immunization, and the influence of service delivery systems on people's perceptions and behaviour. |
